# The natural history of CDKL5 deficiency disorder into adulthood

**DOI:** 10.1101/2025.01.12.24318239

**Authors:** Angel Aledo-Serrano, David Lewis-Smith, Helen Leonard, Allan Bayat, Mohamed Junaid, Eveline Hagebeuk, Christina D Fenger, Juliana Laze, Alessandra Rossi, Marina Trivisano, Beatriz Gonzalez-Giraldez, Julio Lama, Ilona Krey, Konrad Platzer, Elise Brischoux-Boucher, Catherine Sarret, Lysa Boissé Lomax, Caterina Zanus, Luciana Musante, Paola Costa, Patrick Moloney, Norman Delanty, Angelo Russo, Bitten Schönewolf-Greulich, Anne-Marie Bisgaard, Colette Berger, Elena Freri, Satoru Takahashi, Pia Zacher, Julien Jung, Scott Demarest, Eric Marsh, Alan Percy, Jeff Neul, Heather Olson, Lindsay Swanson, Stefano Meletti, Maria Cristina Cioclu, Quratulain Z. Ali, Ana Suller, Alvaro Beltran-Corbellini, Antonio Gil-Nagel, Xiaoming Zhang, Roberto Previtali, Anne F Højte, Nicola Specchio, Jenny Downs, Gaetan Lesca, Guido Rubboli, Danielle Andrade, Elena Gardella, Elia Pestana, Orrin Devinsky, Timothy Benke, Ingo Helbig, Rhys Thomas, Rikke S. Møller

**Affiliations:** Epilepsy Unit, Vithas Neuroscience Intitute, Vithas Madrid University Hospitals, Madrid, Spain; FutureNeuro, Royal College of Surgeons in Ireland University of Medicine and Health Sciences, Dublin, Ireland; Telethon Kids Institute, Centre for Child Health Research, The University of Western Australia, Perth, Australia; Pediatric Neurology Department, Danish Epilepsy Center, Denmark; Department of Pediatric Neurology, Stichting Epilepsie Instellingen Nederland, The Netherlands; Department of Epilepsy Genetics and Personalized Medicine, Danish Epilepsy Centre, Dianalund, Denmark; Amplexa Genetics A/S, Odense, Denmark; New York University Langone Medical Center Comprehensive Epilepsy Center, New York; Department of Epilepsy Genetics and Precision Medicine, Danish Epilepsy Center Filadelfia, member of the European Reference Network EpiCARE, Dianalund, Denmark; Pediatric Neurology Department, Bambino Gesu University Hospital, Rome, Italy; Epilepsy Unit, Fundación Jimenez Diaz University Hospital, Madrid, Spain; Pediatric Neurology Department, Torrecardenas Hospital, Almeria, Spain; Institute of Human Genetics, University of Leipzig Medical Center, Leipzig, Germany; Centre de genetique humaine, CHRU Besancon, Universite de Franche-Comte, Besancon; CHU Clermont-Ferrand, France; Queen’s University, Kingston, Canada; Child Neuropsychiatry Unit, Institute for Maternal and Child Health –IRCCS “Burlo Garofolo”– Trieste; Department of Clinical and Experimental Epilepsy, UCL Queen Square Institute of Neurology, London, UK; Department of Neurology, Beaumont Hospital, Dublin, Ireland and FutureNeuro, the Science Foundation Ireland Research Centre for Chronic and Rare Neurological Diseases, Royal College of Surgeons in Ireland, Dublin, Ireland; Department of Pediatric Neurology, Istituto delle Scienze Neurologiche di Bologna, Bologna, Italy; Department of Clinical Genetics, Copenhagen University Hospital Rigshospitalet, Copenhagen, Denmark; Department of Pediatrics and Adolescent Medicine, Center for Rett Syndrome, Copenhagen University Hospital, Rigshospitalet, Copenhagen, Denmark; Department of Neurology, University Hospitals of Lyon (HCL), Member of ERN EpiCARE, Lyon, France; Department of Pediatric Neuroscience, Fondazione IRCCS Istituto Neurologico Carlo Besta, Milan, Italy; Department of Pediatrics, Asahikawa Medical University, Asahikawa, Japan; Epilepsy Center Kleinwachau, 23. Department of Neurology, University Hospitals of Lyon (HCL), Member of ERN EpiCARE, Lyon, France; Department of Pediatrics and Neurology, Precision Medicine Institute, University of Colorado School of Medicine and Children’s Hospital Colorado, Aurora, Colorado, USA; Division of Child Neurology, Children’s Hospital of Philadelphia, Department of Neurology, University of Pennsylvania Perelman School of Medicine, Philadelphia, Pennsylvania, USA; University of Alabama at Birmingham Heersink School of Medicine, Birmingham, AL, USA; Vanderbilt University Medical Center, Nashville, TN, USA; Department of Neurology, Boston Children’s Hospital, Boston, Massachusetts, USA; Neurology Unit, OCB Hospital, AOU Modena, Modena, Italy; Adult Genetic Epilepsy (AGE) Program, Krembil Brain Institute, Toronto Western Hospital, University Health Network, University of Toronto, Toronto, Ontario, Canada; Department of Clinical Neurological Sciences, Schulich School of Medicine and Dentistry, Western University, London, Ontario, Canada; Epilepsy Program, Neurology Department, Ruber Internacional Hospital, Madrid, Spain; Epilepsy Center, Neurological Institute, Cleveland Clinic, Cleveland, OH, USA; Pediatric Neurology, Department of Biomedical and Clinical Sciences, Buzzi Children’s Hospital, University of Milan, Milan, Italy; Telethon Kids Institute, Centre for Child Health Research, The University of Western Australia, Perth, Western Australia, Australia; Division of Neurology, Children’s Hospital of Philadelphia, Philadelphia, USA; Department of Biomedical and Health Informatics (DBHi), Children’s Hospital of Philadelphia, Philadelphia, USA; Translational and Clinical Research Institute, Newcastle University, Newcastle upon Tyne, UK

**Keywords:** CDD, genetic epilepsy, Rett, developmental and epileptic encephalopathy, neurogenetics

## Abstract

Knowledge of the natural history of *CDKL5* deficiency disorder (CDD) is limited to the results of cross-sectional analysis of largely pediatric cohorts. Assessment of outcomes in adulthood is critical for clinical decision-making and future precision medicine approaches but is challenging because of the diagnostic gap and duration of follow-up that would be required for prospective studies. We aimed to delineate the natural history retrospectively from adulthood.

We analyzed clinical data about an international cohort of 67 adults with CDD. We analyzed demographic, phenotypic, CDKL5 Developmental Score (CDS), and treatment data, and tested associations with genetic factors, sex, and a positive or negative history of neonatal seizures, as an early predictor of prognosis.

All but one of 67 adults (55 females, median age of 24 years at last follow-up) had epilepsy, typically beginning with epileptic spasms or tonic seizures before 4 months of age. Focal-onset and non-motor seizures emerged later. Fewer than a third had been documented as having bilateral tonic-clonic seizures or status epilepticus. Seizures often improved with age, but 73% had never experienced more than 6 months of seizure-freedom. Clobazam, sodium valproate, and lamotrigine were the most frequently prescribed antiseizure medications, but no specific treatment demonstrated superiority. Common comorbidities included movement disorders, visual impairment, sleep disorders, constipation, and scoliosis.

All participants had intellectual disability, 75% had not acquired speech and 45% had regressed developmentally. 16% never achieved any CDS skill, but most attained at least three, and 28% attained six or all seven. By adulthood, half of those who had achieved any CDS skill retained all their CDS skills. The skills most frequently lost were independent walking and standing.

Those with a history of neonatal seizures tended to attain fewer CDS skills and were more likely to have abnormal muscle tone in adulthood, atrioventricular conduction delay, and potential complications of their illness and treatment.

Individuals carrying missense variants attained more CDS skills than those with other variants and were more likely to lose skills in adulthood and develop anxiety, possibly reflecting the limited neurodevelopment of those with non-missense variants, who manifested a more multisystemic disorder.

In summary, retrospective data from adulthood elucidates the evolution of symptoms, variation in developmental outcomes, and the treatment landscape in CDKL5 deficiency disorder. Presence a non-missense variants or a history of neonatal seizures indicates a more complex disorder and lower developmental trajectory. Our findings will inform management decisions, prognostication, and the design of clinical trials in CDKL5 Deficiency Disorder.

## Introduction

Cyclin-dependent kinase-like 5 (CDKL5), encoded by *CDKL5* (Xp22.13), is a serine/threonine kinase with multiple phosphorylation targets. Its deficiency results in abnormalities of axonal and dendritic development and morphology, with impaired synaptic vesicle recycling (reviewed by Van Bergen *et al.*^1^). Pathogenic variants in *CDKL5* cause one of the more common genetic developmental and epileptic encephalopathies (DEEs); CDKL5 deficiency disorder (CDD), which presents with seizures before 36 months of age in 2.36 per 100,000 livebirths.^2,3^

CDD is an early-onset severe neurodevelopmental disorder with drug-resistant seizures, motor and cognitive developmental stagnation or regression, behavioral disturbances, cerebral visual impairment, sleep dysregulation, and gastrointestinal dysfunction.^4,5^ Accurate and precise prognostication in clinic requires comprehensive knowledge of the clinical landscape and range of trajectories of a disorder, which could be enhanced for a particular patient according to known associations between clinical features, or genotype-phenotype associations. In other DEEs some comorbidities become apparent in adulthood.^6–9^ Consequently, knowledge of the natural history of disorders is necessary for clinicians to prioritize potential comorbidities for screening during follow-up and to benchmark for the evaluation of potentially therapeutic interventions. However, no studies have focused on the clinical evolution of CDD through to, and within, adulthood, which limits the evidence available to inform management of adolescents and adults and to prognosticate. Full evaluation of the efficacy of candidate disease-modifying treatments will require greater elucidation of the natural history.

Previously, a cross-sectional study of two composite outcome measures in 285 people with CDD found associations with some recurrent genotypes.^4^ However, the median age was 4.3 years, and the number of adults was unclear: only 38 participants had been followed up beyond 13 years of age. Here, we report the clinical features and histories of 67 adults with disease-causing *CDKL5* variants, describing how the clinical manifestations of CDD evolved into adulthood, and assessing the effect of sex, genotype, and onset with neonatal seizures.

## Materials and Methods

### Protocol approval, participant consent, recruitment, and data collection

Written informed consent for participation/publication was obtained from all parents or legal guardians of participants. This retrospective study protocol and design were approved by the ethics committee of Madrid Ruber International Hospital and the New York University Langone Health IRB. We have sought compliance with the Strengthening the Reporting of Observational studies in Epidemiology (STROBE) guidelines for a cross-sectional study.^10^ A methodological summary follows with full details in the **Supplementary Methods**. Code is available at https://github.com/DLewisSmith/CDKL5_adults. Deidentified data can be shared by request to AAS.

We recruited participants through an international network of pediatric and adult neurologists and medical geneticists, and the International CDKL5 Disorder Database (ICDD).^11^ For inclusion, participants had to be at least 18 years old at last clinical review and have a pathogenic or likely pathogenic variant^12^ of *CDKL5* (MANE select transcript^13^: NM_001323289.2, ENST00000623535.2), assessed using *InterVar*.^14^ We obtained updated information about individuals who have been reported previously,^15–19^ contacting caregivers of the 34 participants from the ICDD where necessary to clarify details they had provided.^11,20–29^

### Mapping of phenotypic data

Clinical data were entered into a study-specific REDCap repository^30^ we adapted from the Epi25 Collaborative REDCap template.^31^ The REDCap repository is hosted at the Danish Epilepsy Centre, Filadelfia. We mapped clinical features from REDCap fields to Human Phenotype Ontology (HPO) concepts (version 1.7.14, release date 2022-02-14) with binning of quantitative gestational age, birth weight, and head circumference data and modification of relationships between concepts as described previously.^32^ We added 19 study-specific concepts to the HPO to maximize the capture of clinical features that have been prioritized in CDD (using the in-house concept unique identifier *mHP*, **Supplementary Table 2**). We annotated each person with those phenotypes asserted to be present (*positive annotations*, using the standard concept unique identifier *HP*) with the age of onset and last occurrence where available, and those asserted to be absent up until the age of data collection (*negated annotations*, modifying the concept unique identifier to *NP* or *mNP*).^33^

### Clinical outcomes

We explored the onset and longitudinal duration of each seizure type in each person by propagating their positive seizure annotations with the annotations of the earliest and latest recorded seizure of that type, and then taking the minimum and maximum age of each seizure type.

Epilepsy outcomes and the consequences of antiseizure medication (ASM), vagus nerve simulation (VNS), and the ketogenic diet were annotated based on the impressions of caregivers and treating physicians. Current seizure freedom was defined as the period without seizures being at least three-fold longer than the longest inter-seizure interval in the preceding year,^34^ with a minimum of 6 months to overlook any transient honey-moon period.^35,36^

CDKL5 Developmental Score (CDS) was calculated on a scale of 0–7 by summation of developmental skills obtained.^18^ We aimed to identify the developmental milestones eventually acquired by adulthood and how many people had lost skills by their most recent follow-up. Precise age annotations of CDS were rarely available. However, we were able to calculate the person’s CDS total from item scores at most recent follow-up in adulthood (when clinically stable) and compare this to their historic maximum to allow identification of skill loss. A participant’s maximum CDS we refer to is age-agnostic. Consequently, a person may have never held all the skills concurrently.

### Stratification

We undertook analyses after stratifying people demographically, genetically (primarily by the coarse dichotomy between missense and non-missense *CDKL5* variants because of the uncertainty as to the optimal stratification by more complex features),^4,11^ and by the presence or absence of neonatal seizures (an early onset phenotype that may be the first manifestation of an underlying severe neurodevelopmental disorder, and/or may contribute to the underlying disorder as an epileptic encephalopathy)^37^, hypothesizing that these discernable features might predict different clinical trajectories.

### Statistical analysis

Common statistical tests are described in the results. All analyses were conducted using the R statistical language.

We quantified the association between the presence and absence of an HPO annotation with membership of these strata, testing positive and negated phenotypes separately (for example, one contingency table for each of *Neonatal electro-clinical seizure [HP:0032809]* and *no Neonatal electro-clinical seizure [NP:0032809]*) in recognition of phenotyping gaps.^38^ We report the conditional maximum likelihood estimate derived odds ratios (OR), relative risks (RR), and unadjusted two-sided *p*-values using Fisher’s exact test (FET, stats::fisher.text()), and applied the *minP stepdown* procedure (*minPsd*, NRejections::adj_Wstep() version 1.2.0) using 100,000 simulations to each stratum.^32,39^

We used phenotypic similarity analysis to quantify the clinical similarity of people in the same group across the combination of all their positively annotated phenotypes, in the context of the cohort, interpreted using a Monte Carlo simulation of 1,000,000 simulations.^38^ In addition to 25 previously established algorithms, we introduce a new minimal term overlap similarity algorithm (*Sim_MinTO_*) as our primary measure. *Sim_MinTO_*quantifies the similarity of two people based on the monotonically modified information content of the HPO concepts sufficient to infer the intersection of the full sets phenotypic descriptors annotated as present in both of the pair. This models how a clinician might summarize the similarity of two people by listing only the descriptors necessary to capture all the pair’s shared clinical features as succinctly as possible and considering how informative these are in a context-dependent manner. It takes an open-world interpretation that acknowledges phenotyping gaps in real-word data.

## Results

### Description of the study cohort and variants

Sixty-seven adults from 66 families (one pair of monozygotic twins) were recruited across 13 countries, 20 from the ICDD (**Figure 1A**, **Table 1, Supplementary Table 1**). The specialty of the most recent responsible neurologist was known for 46, with the 26 (57%) remaining under the care of a pediatric neurologist tending to be younger than the 20 (42%) under an adult neurologist (median age at last follow-up=21.5 versus 28.0 years old, range=18–36 versus 18-57 years old, Wilcoxon rank sum test with continuity correction, WRST W =148, two-sided *p*=0.0130). Beyond large deletions (in ten people), 49 different variants were detected (**Figure 1B**).

**Figure 1.**
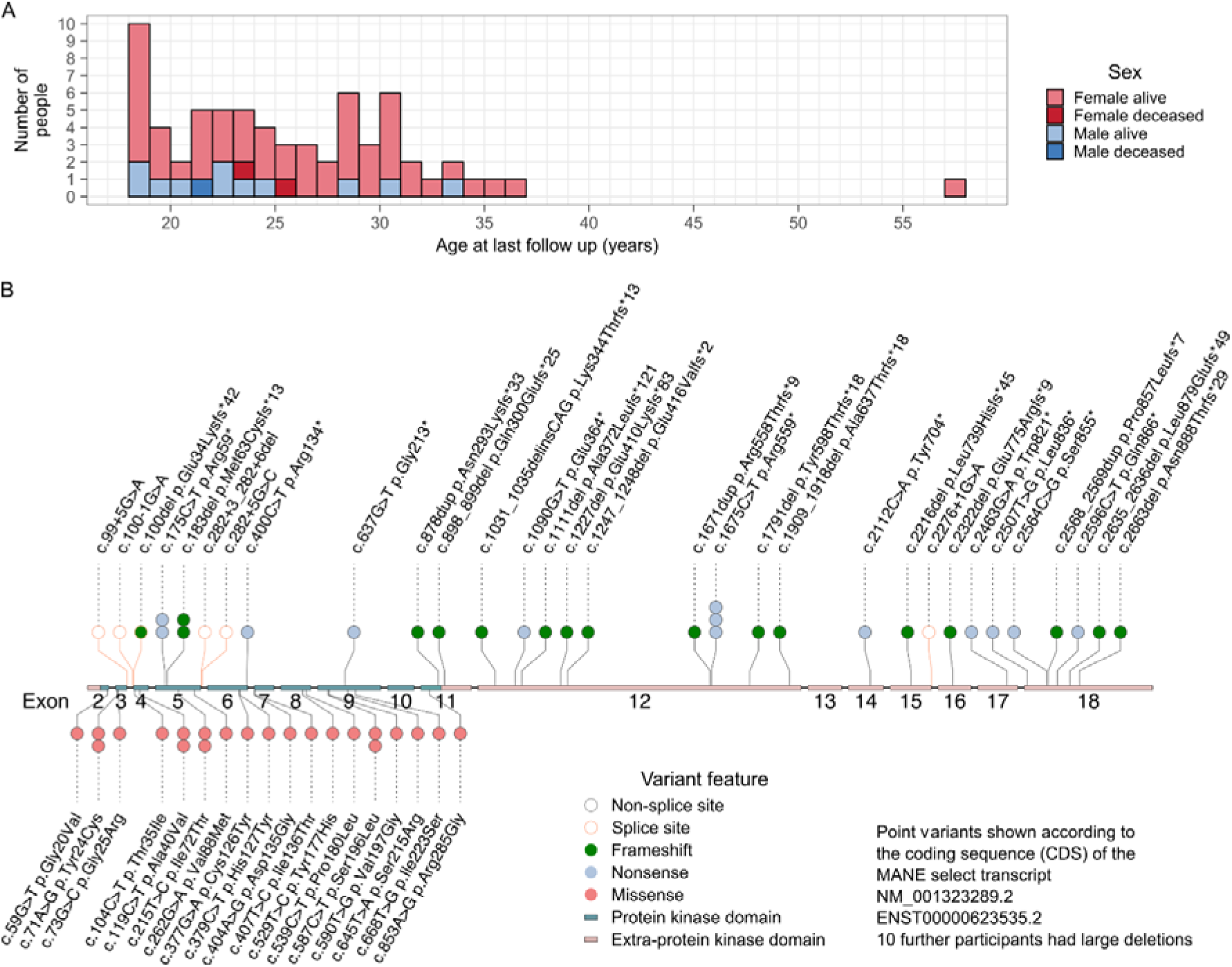
Age at last follow-up (A) and *CDKL5* variants (B) in the cohort. c.183del (p.Met63Cysfs*13) was carried by a pair of monozygotic twins.

**Table 1.** Description of the demographics, CDKL5 Developmental Scores, and variants of the cohort. ^a^ Minimum total CDKL5 Developmental Score=0, maximum=7. ^b^ Among those attained the skill.

| Feature | Number or median<br>(Number with data) | Percentage of those with data or<br>minimum, interquartile range,<br>maximum |
| --- | --- | --- |
| **Sex** |  |  |
| Female | 55 (67) | 82% |
| Male | 12 (67) | 18% |
| Vital status |  |  |
| Alive at last data collection | 64 (67) | 96% |
| Deceased | 3 (67) | 4.5% |
| Accommodation at last follow-up |  |  |
| Most nights spent with family | 45 (66) | 68% |
| Completely institutionalized | 15 (66) | 23% |
| Lead neurologist at last follow-up |  |  |
| Pediatric | 26 (46) | 57% |
| Adult | 20 (46) | 43% |
| **Variant summary** |  |  |
| Missense kinase domain | 22 (67) | 33% |
| Non-missense kinase domain | 12 (67) | 18% |
| Non-missense non-kinase domain | 23 (67) | 34% |
| Large deletion | 10 (67) | 15% |
| Confirmed de novo | 40 (41) | 98% |
| Confirmed inherited | 1 (41) | 2.4% |
| Confirmed mosaicism | 5 (unknown) | 7.5% (of all 67) |
| **CDKL5 Developmental Score** |  |  |
| Total maximum CDKL5 Developmental Score <sup>a</sup> | 3 (67) | 0, 1–6, 7 |
| Ever able to sit independently | 42 (67) | 63% |
| Ever able to stand independently | 32 (67) | 48% |
| Ever able to walk independently | 28 (67) | 42% |
| Ever able to babble | 42 (67) | 63% |
| Ever able to use a word | 17 (67) | 25% |
| Ever able to use a raking grasp | 47 (67) | 70% |
| Ever able to use a pincer grasp | 18 (67) | 27% |
| Recent Total CDKL5 Developmental Score <sup>a</sup> | 2 (67) | 0, 1–4, 7 |
| Able to sit independently at last data collection | 38 (67) | 57% (90% of those attaining skill) |
| Able to stand independently at last data collection | 19 (67) | 28% (59% of those attaining skill) |
| Able to walk independently at last data collection | 14 (67) | 21% (50% of those attaining skill) |
| Able to babble at last data collection | 36 (67) | 54% (86% of those attaining skill) |
| Able to use a word at last data collection | 13 (67) | 19% (76% of those attaining skill) |
| Able to use a raking grasp at last data collection | 42 (67) | 63% (89% of those attaining skill) |
| Able to use a pincer grasp at last data collection | 14 (67) | 21% (78% of those attaining skill) |
| **Age** |  |  |
| Age at last data collection (years) | 24 (67) | 18, 21–29, 57 |
| Age at diagnosis (years) | 16 (55) | 2, 14–22, 51 |
| Attained independent sitting (months) <sup>b</sup> | 12 (25) | 3, 8–15, 26 |
| Attained use of a word (months) <sup>b</sup> | 19 (8) | 15, 17.5–31.5, 72 |
| Attained independent walking (months) <sup>b</sup> | 30 (17) | 12, 17–60, 108 |
| Regression thought have begun in those with any documented age of regression (months) | 132 (27) | 8, 27–228, 624 |
| Regression thought have begun in those with documented age of regression prior to adulthood (months) | 48 (18) | 8, 18–129, 168 |

### Phenotypic annotation

We were able to infer the number of people reported to have and have not manifested each of 507 phenotypes up to the age of data collection (**Supplementary Results**). Unless a denominator is specified, subsequent percentages correspond to the entire cohort (*n*=67).

### Seizures

All but one person experienced seizures (99%). Four (6.0%) had seizures in the first week of life. Thirteen (19%) had been documented as having had, and 45 (67%) as having not had neonatal seizures (phenotyping gap 13%).

Most people had seizures beginning with motor manifestations (*n*=62, 93%), usually tonic seizures (irrespective of electrographic onset, *n*=51, 76%) or epileptic spasms (*n*=43, 64%). The latter usually started in infancy (*n*=37/43, 86%). Among the 13 people with neonatal seizures, these were predominantly epileptic spasms (*n*=7/13, 54%) and tonic seizures (*n*=5/13, 38%). Myoclonic (*n*=26, 39%) and non-motor (*n*=30, 45%) seizures were common. Non-motor seizures comprised atypical absences in 20/30 (67%) and focal non-motor seizures in 15/30 (50%). Five of 30 (17%) had both. Broadly, seizures could be classified as focal-onset in 28 (42%) and generalized-onset in 20 (30%), with eight (12%) having both. In 26 people (39%) no electroclinical onset was documented. Bilateral tonic-clonic seizures (regardless of onset) were reported in 21 (31%). and seizures with a dialeptic component (incorporating absence and focal impaired awareness seizures) in 25 (37%). Status epilepticus had occurred in 14 (21%), more commonly specified without (*n*=11, 16%) rather than with (*n*=1, 1.5%) prominent motor symptoms. Forty-eight (72%) had not had recognized status epilepticus. Multiphasic seizures with evolving motor components were documented in eight (12%) and described in six, typically involving a tonic phase (*n*=5/6) that evolved into spasms (*n*=4/5).

Onset of seizures was before 2 months of age in half, and before 11 months in all 38 with complete ages of onset for all documented seizure types (**Figure 2**). Repeating the analysis in all 66 with seizures, censoring seizure types with missing age of onset gave an upper limit for the median age of seizure onset in this cohort within the fourth month (cumulative risk of 62% by 4 months old). Fewer than 20% were yet to manifest a seizure at 9 months old.

**Figure 2.**
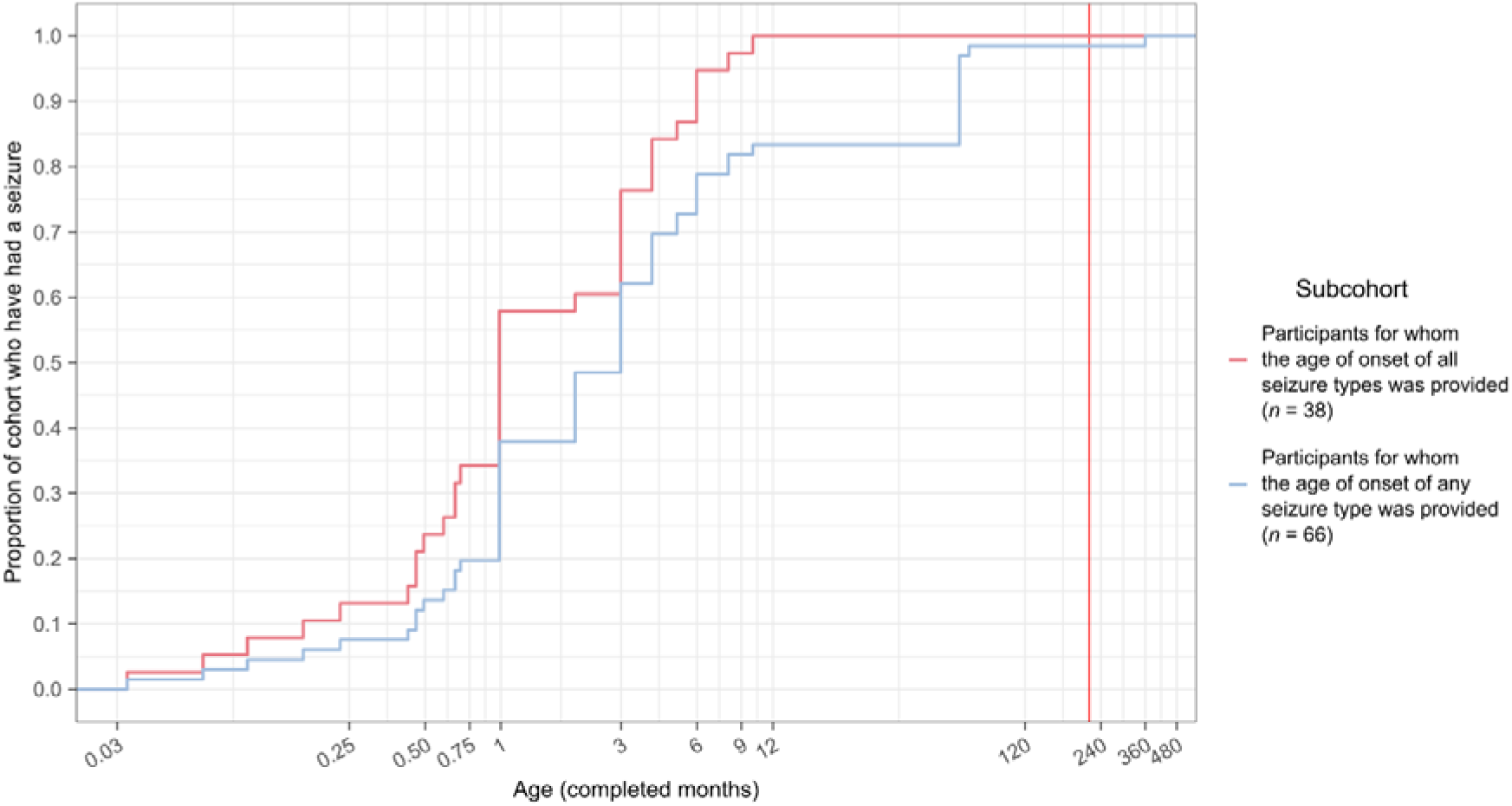
The distribution of age of onset of epileptic seizures of any type. The vertical red line indicates 18 years of age, beyond this age, follow-up is incomplete.

So far, five of 66 people with epilepsy (7.6%) had manifested new types of seizure (myoclonic, focal impaired awareness emotional, and atypical absence seizures) in adulthood, aged 18–23 years (**Supplementary Results**). To evaluate the chronological pattern of seizure types in CDD, we summarized the age of onset of each seizure type in the people with sufficient data (**Figure 3A**). Tonic seizures and epileptic spasms dominated infancy. Tonic-clonic, focal-onset, myoclonic, and non-motor seizures began in early childhood, with seizures that have initial motor semiology tending to present earlier than non-motor seizures. Atonic seizures and status epilepticus, usually non-convulsive, presented in later childhood. Twenty-five people had been annotated with at least one seizure type complete with not only an age of onset but also the age of most recent occurrence. These limited data suggest that epileptic spasms entered long-term remission in most, but not all, by adolescence (**Supplementary Results**).

**Figure 3.**
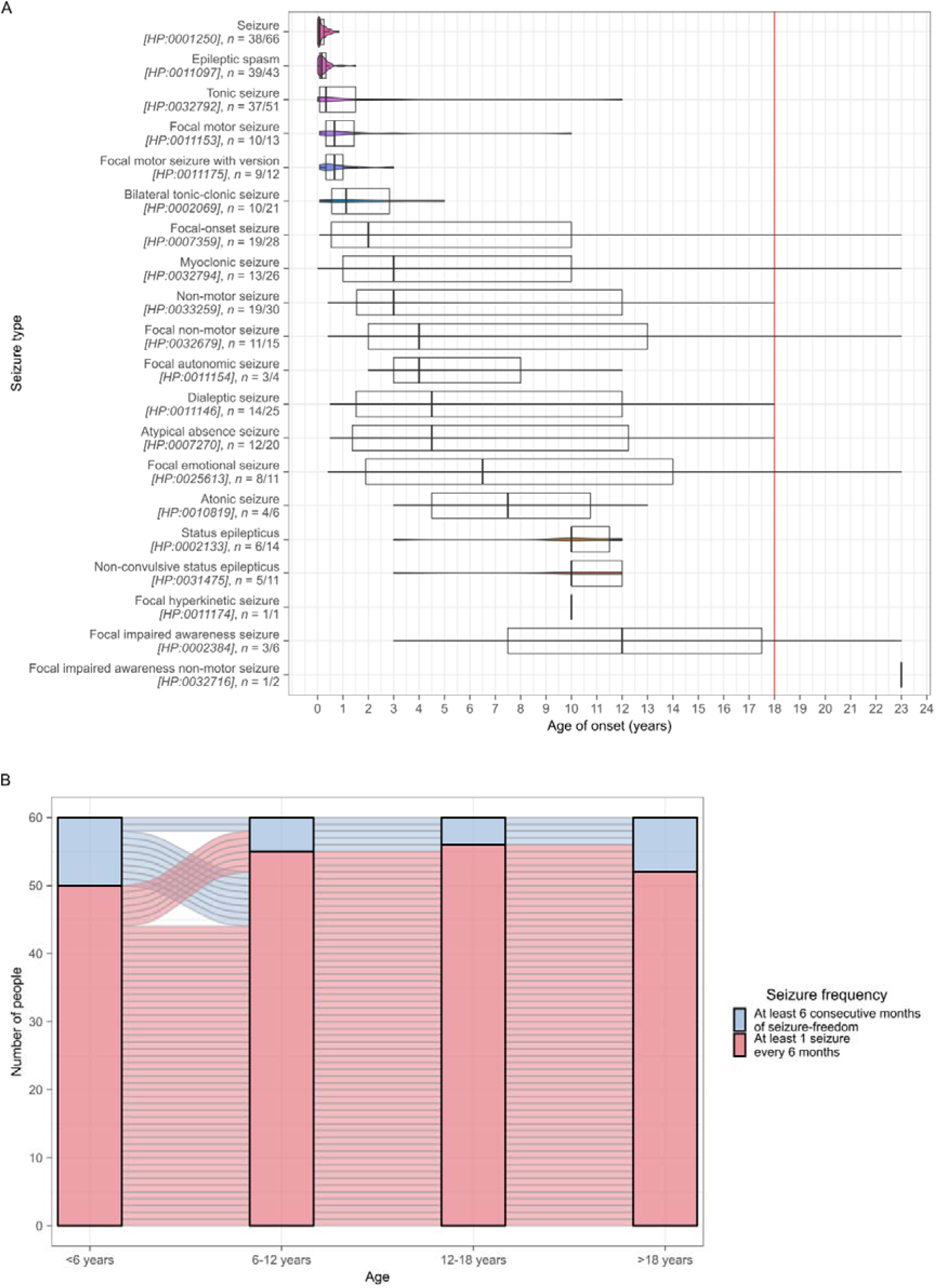
The evolution of seizure types by onset (A), and 6-year intervals containing 6 months of seizure freedom (B).For 60 people with epilepsy, we could assess the occurrence of six consecutive months of seizure-freedom in each consecutive 6-year period from birth to adulthood (Figure 3B). Fewer than 20% attained 6 months of seizure-freedom in any 6-year period, the majority (*n*=44, 73%) never did, and only two (3%) attained it consistently.

At last data collection, of 62 adults who remained alive with a history of seizures (data missing for one), 35 (56%) had seizures at least daily, a further ten (16%) at least weekly, and nine (15%) at least monthly. Eight (13%) had been seizure-free for at least 6 months: 5/8 (62.5%) continuing and 3/8 (37.5%) having stopped ASM. Catamenial worsening of seizures was reported for 11/55 women (20%), denied for two (3.6%), and unknown in 42 (76%).

A caregiver’s or physician’s subjective description of the evolution of childhood-onset seizures in adulthood was available for 63 (95% of 66 with seizures). Twenty-six (41%) had an improvement (eight, 13% seizure-free), 31 (49%) remained stable, and seven (11%) had worsening (exact binomial test two-sided *p*=00132 for equal probabilities of improvement and worsening among 33 reporting a change in seizures). The oldest individual (range of age between 55-60 old) had periods of improvement and worsening (included in both figures). Periods of improvement or worsening were thought to have occurred in response to treatment changes in 13 and two, and spontaneously in eight and four, respectively. While the proportions favored changing treatment, this did not reach significance (13/8 versus 2/4, relative risk, RR=3.3, OR=3.1, 95% CI=0.35–42, FET unadjusted two-sided *p*=0.358).

### Electroencephalography

Electroencephalographic abnormalities had been documented in 46 people (no reports for 21). Most had generalized slow activity. Thirty-eight (57%) had epileptiform discharges: usually focal (*n*=35/38, 92%), frequently multifocal (*n*=15/35, 43%). Generalized discharges were reported in 22/38 (58%), most of whom also had focal epileptiform discharges on the same or another electroencephalogram (*n*=19/22, 86%). Hypsarrhythmia and a photoparoxysmal response were documented in one individual each. Electroencephalographic reports in adulthood and those in childhood differed only in the provision of spatial localization of focal epileptiform discharges in adulthood (**Supplementary Results**).

### Antiseizure treatment

The 66 people with epilepsy had been exposed to 36 different antiseizure treatments (**Supplementary Results**). The median number of antiseizure treatments to which a participant had been exposed was seven (mean*=*7.7, range=1–18). Of the total 513 treatment exposures, 40 were considered to have benefitted (7.8%) and 25 to have aggravated seizure control (4.9%) without specifying seizure type. At most recent follow-up, 3/66 (4.5%) were seizure-free without continuing ASM. In the remaining 63, the median number of regular antiseizure treatments was three (mean*=*2.95, range=1–6), with four people (6.3%) additionally prescribed a rescue medication. More than half had been exposed to lamotrigine (59%), clobazam (58%), sodium valproate (56%), levetiracetam (55%), and topiramate (52%). The most commonly prescribed currently (among those 63 receiving ASM at last follow-up) were clobazam (40%), sodium valproate (32%), and lamotrigine (25%), with 41/63 (65%) taking at least one, 17 (27%) at least two, and three (4.8%) taking all three of these.

We could not demonstrate that any particular treatment was robustly beneficial or detrimental to seizure control after Holm’s adjustment for multiple testing. Ketogenic diets, attempted by 21 people (two in adulthood), were considered beneficial in one and thought to have aggravated seizures in another. All had ceased the diet by last follow-up. VNS was started in 19 (five in adulthood) and continued in ten to last follow-up, one of whom was seizure-free. It was considered beneficial in three and aggravating in none.

Fifty-three courses of treatment were commenced in adulthood involving 21 treatments in 22 adults (median=22, range=1–7 among these, **Supplementary Results**). Sixteen (30%) were considered beneficial, and two (3.8%) were considered detrimental (topiramate and primidone in different people).

### Neurodevelopmental, psychiatric, and other neurological features

All had intellectual disability, usually profound or severe (*n*=61, 91%). Fifty (75%) had not acquired speech. Neurodevelopmental delay was reported in 54 (81%), noted prior to the onset of seizures in 18/54 (33%), and usually specified as gross motor (*n*=39/54, 72%) or global (*n*=18/54, 33%). Fine motor delay was annotated to only a subset of those with gross motor delay (*n*=20/39, 51%). Developmental regression was documented thoroughly, occurring in 30 (45%) and not in 36 (54%). Regression occurred during periods of subjectively higher (*n*=14/30, 47%) and lower (*n*=14/30, 47%) seizure burden, including in one person when seizure-free. Twelve of 30 (40%) lost skills in adulthood. The person without seizures had not regressed.

Autistic behavior was reported in 15 (22%). Attention deficit hyperactivity disorder was reported in three (4.5%), improving during adulthood in all, and affective disorders in seven (10%): five with anxiety and three with depression, including one with both. Psychosis was not reported. Self-injurious behaviors (all self-biting) were documented in 18 (27%), and other aggressive behaviors in 28 (42%). Aggressive behaviors began in adulthood in seven of 28 (25%). In those 21 with aggressive behaviors beginning before adulthood, behavior improved in five (24%) in adulthood, remained stable in eight (38%), and worsened in eight (38%).

Stereotypies were reported in 49 people (73%), most frequently hand wringing (*n*=42/49, 86%), a tendency to insert their hand in their mouth (*n*=29/49, 59%), or body rocking (*n*=15/49, 31%). All 18 people with self-injurious behavior had stereotypies.

Sixty-two people (93%) were documented as having either *Abnormality of movement [HP:0100022]* or *Abnormality of central motor function [HP:0011442]*. Both abnormalities of coordination (*n*=52, 78%) and involuntary movements including stereotypies (*n*=51, 76%) were common. Beyond stereotypies, specific hyperkinetic disorders occurred in 18 (27%), including dystonia in 11 (16%); or the chorea-dyskinesia spectrum in nine (13%); and tremor in two (3%). Hypokinesia (*n*=6, 9.0%) and bruxism (*n*=4, 6.0%) were reported more rarely. Mobility problems were reported in 51 (76%) and included gait disturbance (*n*=47, 70%), often described as shuffling (*n*=5/47, 11%) or ataxic (*n*=3/47, 6.4%), and postural instability (*n*=13, 19%). Thirty-nine never walked (58% of the cohort, 76% of those 51 with gait disturbance or postural instability).

Sixteen (24%) had a parkinsonian feature (bradykinesia, hypokinesia, postural instability, retrocollis, or short-stepped shuffling gait), usually beginning in adolescence. We did not find an association between greater age at last assessment and either the presence of a parkinsonian feature (WRST W=361.5, one-sided *p*=0.756) or the number of parkinsonian features (Kendall’s τ=-0.051, one-sided *p*=0.693). Despite the absence of chronological evidence of causation, four people with parkinsonian features were exposed to valproic acid (*n*=2) or antipsychotics (*n*=2). One person had self-limited focal dystonia following exposure to metoclopramide during adulthood. Medications prescribed for movement disorders included trihexyphenidyl (*n*=2), levodopa (*n*=1), and amantadine (*n*=1), without data on response. The evolution of movement disorders was described in 26. In 16/26 (62%) these began in adulthood. Of the 10 with movement disorders beginning before, eight became more severe and two remained stable in adulthood.

Abnormal muscle tone at some age was reported in 32 (48%). Neonatally, hypotonia was reported in 18 (27%) and excluded in 8 (12%). Neonatal hypertonia was not recorded. In adulthood, 21 (31%) had hypertonia, usually spasticity with one treated with botulinum toxin and another with baclofen. Hypotonia was documented in two adults (3%).

Sleep disorders were reported in 43 (64%), with difficulty with sleep maintenance, sleep-wake circadian rhythm disturbance, and sleep onset insomnia common among these. Sleep apnea was reported in five of 43 (12%). One person had abnormal rapid eye movement sleep, in addition to postural instability, stooped posture, and short stepped shuffling gait. Sleep disorders were adult-onset in ten of 43 (23%). In those with earlier onset sleep disorders, these improved in seven (21%), worsened in two (6.1%), or remained stable in 24 (73%) in adulthood. Medications for sleep symptoms included melatonin (*n*=8), trazodone (*n*=5), benzodiazepines (*n*=3), clonidine (*n*=1), and zopiclone (*n*=1), as well as antidepressants and antipsychotics for which sleep may have contributed to the indication.

Visual impairments, always interpreted as cerebral, were recorded in 44 (66%).

### Neurodevelopmental evolution

We determined each participant’s total maximum and the most recent total CDS at assessment in adulthood (**Table 1**, **Figure 4**). The median age at which children were able to sit was 12 months, to walk was 30 months, and to use a word was 19 months (**Supplementary Results**). The median age at which regression was reported to have begun was 48 months among those 18 demonstrating regression prior to adulthood and 132 months including those with loss of skills during adulthood (**Table 1**, **Supplementary Results**).

**Figure 4.**
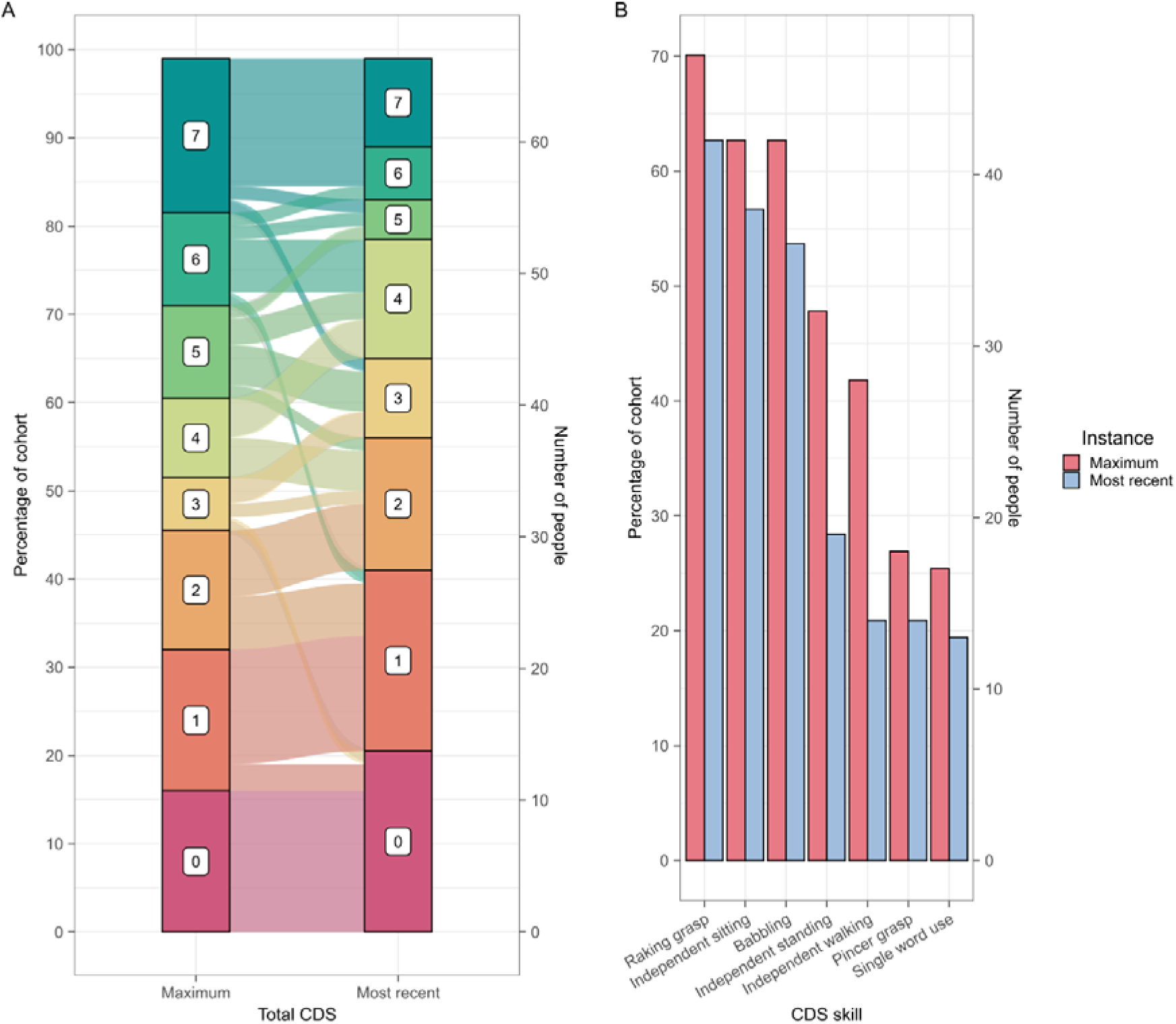
We hypothesized that lower maximum CDS may be associated with early-onset seizures, as a marker of more severe underlying neurodevelopmental encephalopathy, or through the consequences of an early epileptic encephalopathy. The data supported an association with strictly neonatal (WRST *W*=430.5, *p*=0.00481, effect size=0.341, Figure 5A) but not later onset seizures (**Supplementary Results**).

Eleven people (16%) never reached any CDS milestone, most attained at least three, and 19 (28%) attained at least six, of whom 12 (18% of 67) attained all seven skills (**Figure 4A**, **Supplementary Results**). Under half were ever able to stand unaided, but most of those who could also attained independent walking (*n*=28/32, 88%). Fewer than half of those able to babble were ever able to use one word (*n*=17/42, 40%). Similarly, nearly half of those who could use a rake grasp were ever able to use a pincer grasp (*n*=18/47, 38%, (**Figure 4B**, **Table 1)**. At last assessment as an adult, the median total CDS was 2, with 14 people (21%) not being able to perform any of the skills, 4 (6.0%) performing six, and seven (10%) performing all seven. Standing and walking independently were the skills most frequently lost. Half of people who had attained any CDS skill retained all their skills into adulthood (*n*=28/56). The number of skills lost increased with the number of skills attained, except in those attaining all seven.

### Comorbidities

Gastroenterological abnormalities were common (*n*=60, 90%), with most (*n*=57, 85%) having suffered constipation (23/57, 34% needing laxatives long-term. Difficulty feeding or dysphagia was common (*n*=40, 59%). Thirty-one (46%) required gastrostomy feeding, usually before adolescence (*n*=26, 39%). Five of 41 (12%) not requiring a gastrostomy by adolescence had the procedure subsequently. Thirty (45%) had gastroesophageal reflux (*n*=27, 40%) or episodic vomiting (*n*=3, 4.5%). Six were treated chronically with anti-reflux medications and four with antiemetics. Two underwent Nissen fundoplication in adulthood. Three used anti-muscarinics to manage saliva. A third of the cohort had experienced at least one episode of aspiration pneumonia.

Fourteen of 42 (21%) people with scoliosis (63% of the cohort) underwent corrective surgery (seven during adulthood). Osteoporosis was diagnosed in eight (12%), and fractures were reported in five (three fibular, one tibial, and one humeral), four of whom had osteoporosis. Hip dislocation (*n*=3, 4.5%) and contractures (*n*=2, 3.0%) were reported rarely.

Significant cardiovascular abnormalities were reported in eight people (12%), each with a single anomaly. Five (7.5%) had electrocardiographic abnormalities, all detected in adulthood: atrioventricular block (*n*=2, 3.0%), as well as atrial flutter, prolonged QTc, and right bundle branch block (each in one person). The single case of reported syncope was vasovagal. Systemic arterial hypertension was diagnosed in two (3.0%) and atrial septal defect with mitral valve prolapse detected in one.

Hypothyroidism was diagnosed in two people (3.0%), one of whom also had type 2 diabetes mellitus. Five (7.5%) developed nephrolithiasis, all in adulthood following topiramate or ketogenic diet treatment, with one requiring surgical treatment.

### Neuroimaging

Seventeen people (25%) had a documented abnormality of central nervous system morphology. Three (4.5%) had microcephaly and two (3.0%) had macrocephaly. Brain atrophy was detected in 14 (21%), specified as cerebral in three (4.5%) and cerebellar in another three (4.5%). One person had spina bifida with a myelomeningocele and another had a thin corpus callosum.

### Mortality

All three who died carried large deletions (one involving the whole *CDKL5* gene). The cause of death was known for two, both dying after hospitalization for infections with seizures worsening sufficiently to have been judged to contribute to death. There was no significant difference between the total maximum CDS of those three who were known to have died (median*=*2) and those 64 still alive (median*=*3) at last data collection (WRST W=92.5, two-sided *p*=0.927, **Supplementary Results**).

### Molecular diagnosis

The year of molecular diagnosis was available for 55 people (82%) with a median age at diagnosis of 16 years (range=2–51 years) and a trend towards more recent diagnoses being at more advanced ages (**Supplementary Results**). Clinicians reported that molecular diagnosis had altered clinical management for 25 participants (37%) and had not for 26 (39%).

We hypothesized that clinician perception of the benefit of obtaining a molecular diagnosis might be explained by seizure burden and the availability of precision medicines. We found those for whom molecular diagnosis was not felt to have altered management were more likely have had 6-month periods of seizure freedom (OR=4.4, FET two-sided *p*=0.00604). Molecular diagnoses perceived as beneficial to management tended to be more recent (median year=2015 versus 2011, WRST W=368.5, two-sided *p*=0.0225). There was no difference in age at diagnosis between those in whom molecular diagnosis affected management and those in whom it had not (medians 16 years and 17 years respectively, WRST W=262.5, two-sided *p*=0.974).

### Phenotypic associations

We dichotomized the cohort into those 22 participants with missense variants and those 45 with other variants and tested the association between these and all 525 informative clinical phenotypes annotated to the cohort (6,376 annotations regarding the presence of 496 and absence of 29 HPO concepts). No missense versus non-missense association with phenotype reached *minPsd* < 0.05 (**Figure 6**, **Supplementary Results**). However, *Regression in adulthood [mHP:0000010]* (OR=5.7, *p*=0.0140) and *Anxiety [HP:0000739]* (OR=9.4, *p*=0.0368) were more common among the 22 people with missense variants. Conversely, morphological, motor, gastrointestinal, infective, and neurodevelopmental abnormalities were more common among the 45 with non-missense variants. People with missense variants tended to attain more CDS skills than those with non-missense variants (**Figure 5B**). Each of these two genetically-defined groups was more phenotypically similar than expected by chance when individuals are compared across all their clinical features (**Supplementary Results**). Phenotypic associations with specific genetic variants, sex and neonatal seizures are included in the **Supplementary Results**.

**Figure 5.**
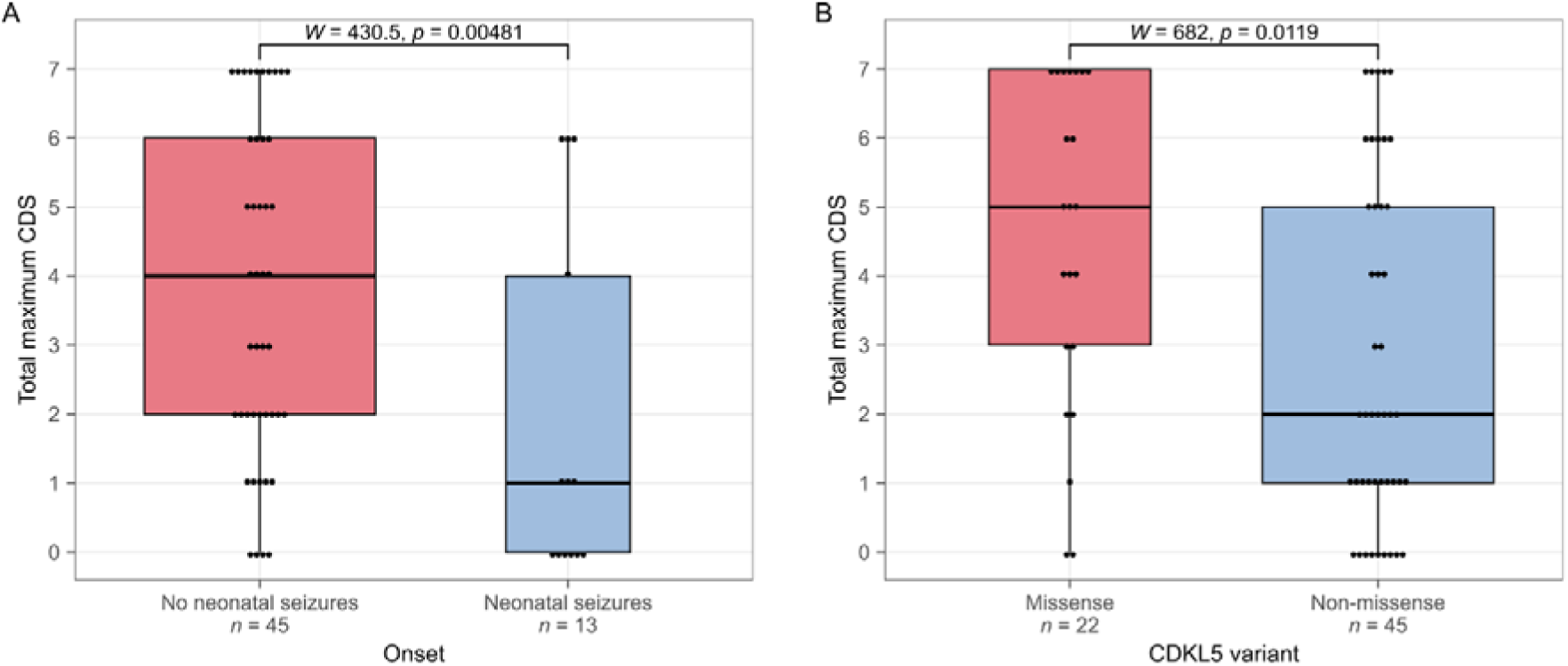

**Figure 6.**
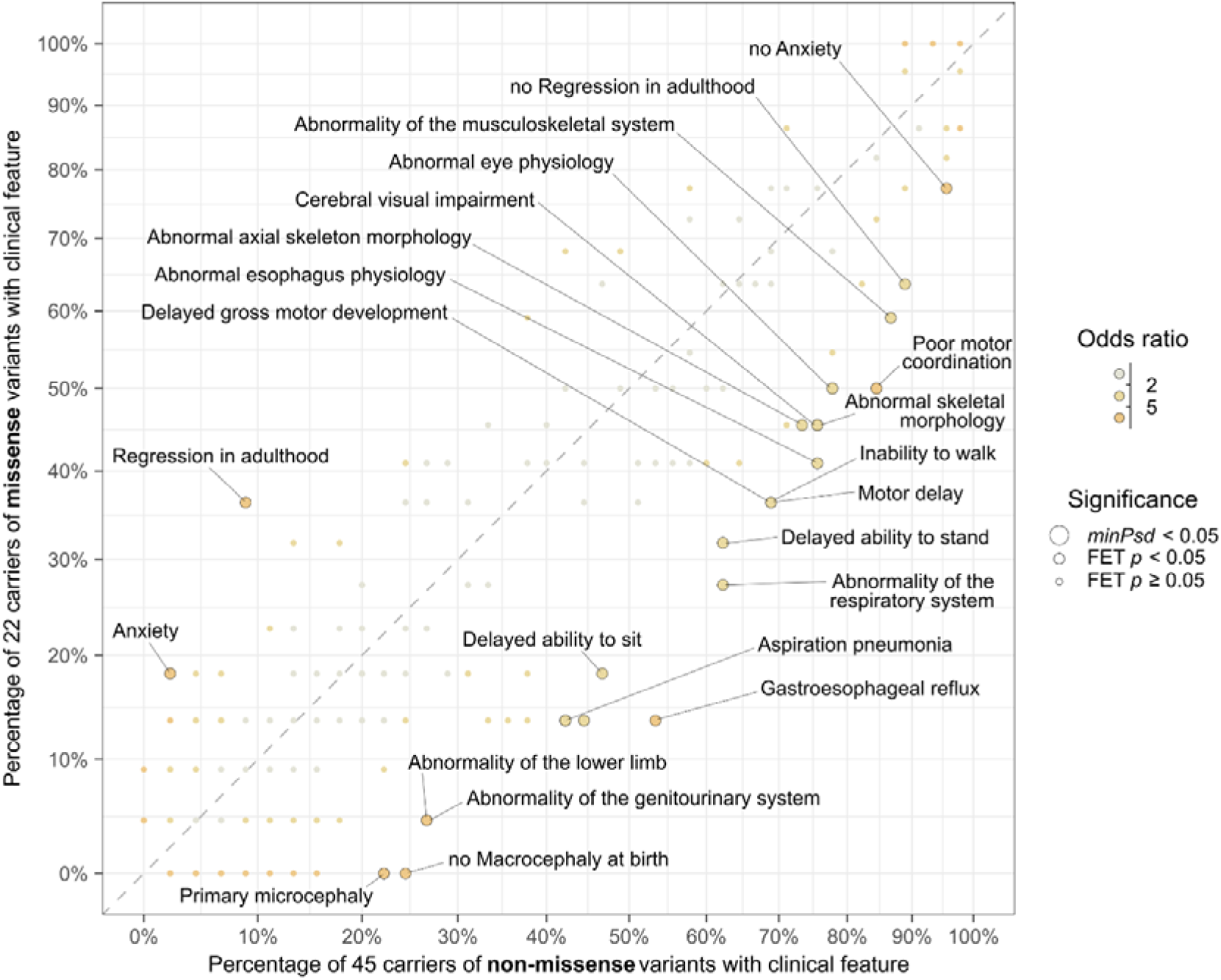

### Predicting neurodevelopmental outcomes

A history of neonatal seizures and whether the *CDKL5* variant was missense or not were independently associated with total maximum CDS (**Supplementary Results**). Consequently, we explored the effect of stratification according to the combination of these two on total maximum CDS, focusing on the 58 people for whom a history of neonatal seizures had been annotated or asserted to have been excluded.

We found significant group-wise differences in total maximum CDS (Kruskal-Wallis rank sum test, *stats::kruskal.test()*, chi-squared=9.85 with 3 degrees of freedom, *p*=0.0199). Those who had both a history of neonatal seizures and a non-missense variant tended to have more limited skill acquisition (median*=*0, mode=0, **Figure 7**). More skills tended to be acquired by those with missense variants and no neonatal seizures (median*=*4–5, mode=7) and by those with non-missense variants and no neonatal seizures (median*=*3, mode=2). Only three people were known to have a missense variant and history of neonatal seizures (median*=*4 range=1–6), which limits the power to interpret comparisons with this combination of risk factors. Importantly for advising patients and caregivers at the individual clinical level, all groups spanned a wide range of total maximum CDS, indicating that regardless of variant type and the history of neonatal seizures, some people with CDD acquire multiple skills, and outcomes must be influenced by other factors. Two with a history of neonatal seizures and carrying non-missense variants attained all skills except a pincer grasp.

**Figure 7.**
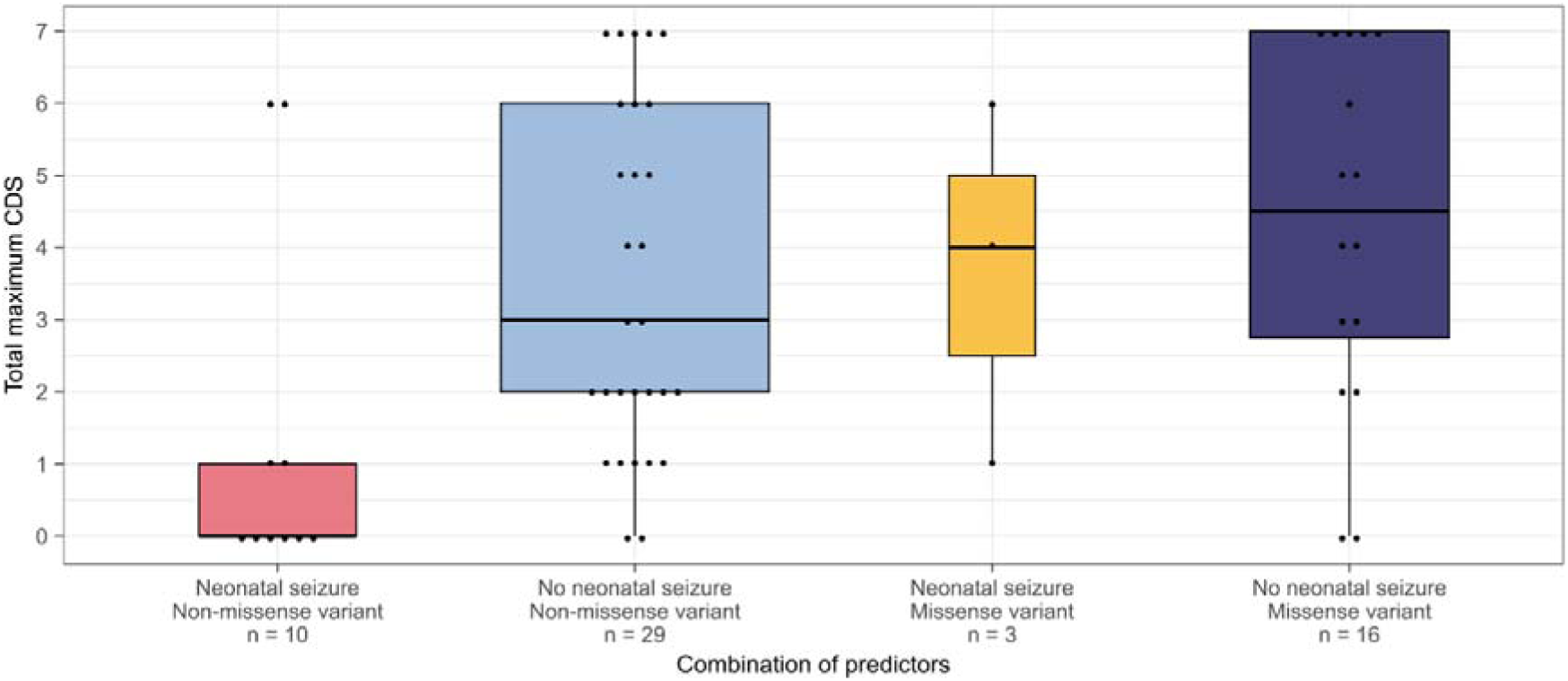
The total maximum CDS by the occurrence of neonatal seizures and whether the diagnostic *CDKL5* variant is missense.

Consider the situation where a genetic diagnosis is obtained after knowing that the person has had neonatal seizures. Without knowing the variant type, we would predict the median total maximum CDS achieved by adulthood in this scenario to be 1 (range 0–6, *n*=13, **Figure 5A**). Finding a missense diagnostic variant would allow revision of this prediction to a median of 4 (range 1–6, *n*=3), and finding a non-missense variant to 0 (range 0–6, *n*=10, **Figure 7**). The variant classification-agnostic median total maximum CDS was 4 (range 0-7, *n*=45) for those without neonatal seizures, with little change on finding a missense (median*=*4–5, range 0–7, *n*=16) or non-missense (median*=*3, range 0–7, *n*=29) variant.

For reference, we provide the proportion of people acquiring each CDS skill across the entire cohort according to sex, history of neonatal seizures, broad variant class, and the combination of the last two of these factors (**Supplementary Results**).

## Discussion

Here, we focus on the clinical features and experiences of *CDKL5* Deficiency Disorders in adults, describing the developmental outcomes and providing a perspective of the range trajectories through to adulthood. Even though it remains possible that this cohort overrepresents early-onset and severe presentations as these features may increase the likelihood of being offered testing and referral to academic centers,^40,41^ we believe that our cohort is representative of the clinical spectrum of CDD in adults. Our retrospective international perspective confirms that epilepsy is common, usually presenting in infancy, and usually remains drug-resistant.^35,42–44^ We confirm that neurodevelopment is delayed with half manifesting regression,^11,20^ and that skill acquisition remains limited despite prolonged follow-up. A history of neonatal seizures and classification of the variant may refine early prognostication.

In our study, we have extended the characterization of seizure trajectory into adulthood, ^18,42^ which was largely unknown to date. Tonic seizures and spasms, predominating in infancy, can persist into adulthood. Myoclonic, tonic-clonic, and focal-onset seizures may emerge from early childhood, with non-motor seizures later. Status epilepticus is rare compared to other genetic DEEs and typically non-convulsive.^45^ The majority of people never obtain 6 months of seizure freedom despite follow-up to adulthood, and most continue to have seizures daily, as in childhood.^35^ However, many have subjective improvement in seizure burden in adulthood and worsening appears rare. EEG changes remain focal or multifocal^42^ and hypsarrhythmia may be rare despite epileptic spasms.^18^

Few adults with CDD were seizure-free without treatment in adulthood and continuing polytherapy was common. We had only pragmatic subjective measures of efficacy, covering only a small proportion of treatment trials. Many treatment regimen settle on clobazam, sodium valproate, or lamotrigine in adulthood, ASM found beneficial in younger individuals.^36^ Newer treatments appear promising. We found little evidence that VNS or ketogenic diets offer long-term benefit in adults. Some people seem to benefit from therapeutic changes in adulthood, although recency and honeymoon periods could have biased evaluation of these most recent changes.^35,36^ Future studies of real-world treatment response may benefit from contemporaneous epilepsy-tailored electronic health records within epilepsy learning healthcare systems.^46^

In CDD, developmental delay can present before seizures, despite their infantile-onset. However, we find that regression is widespread beyond childhood: nearly half lose skills, sometimes in early adulthood and despite improving seizures. We have referred to loss of developmental skills as *regression* pragmatically including when referring to adolescents and young adults because we use it in clinic for want of a term specific to describe the loss of skills in young adults. However, we accept that the loss of skills in early adulthood may be different from that during childhood.^47^

Intellectual disability without speech, with visual impairment and inability to stand or walk independently is typical of CDD in adulthood. One in six adults has never attained any CDS skill, but of those who do, half retain all their CDS skills with 16% performing all or all but one of the skills, and our data do not suggest that skills are gradually lost in early adulthood. Extended follow-up shows that the proportion eventually able to stand and walk independently may be greater than previously found,^11^ but half of those learning to stand or walk will lose this ability. Furthermore, a history of neonatal seizures, present in 19%, predicts more limited developmental outcomes. Whether these contribute to the developmental disorder or indicate manifestation of more severe disease remains unclear, although in pediatric populations low seizure burden may be associated with improvements in CDS.^29^

Movement, gastrointestinal, and sleep disorders remain common in early adulthood, and are occasionally parkinsonian. Many adults use laxatives and may have had gastrostomy or scoliosis surgery. We suspect that our data may underestimate osteoporosis as half of these 12% had suffered fractures. The proportion who had undergone bone dual-energy x-ray absorptiometry is unknown, leading us to suspect that latent osteoporosis may be more common. Finding electrocardiographic abnormalities in 7.5%, is in keeping with findings of prolonged QT_c_ ^48^ and supraventricular tachycardia^49^ in pediatric CDD. These, the osteoporosis, and nephrolithiasis, emphasize the importance of considering the risks of ASMs and screening for subclinical comorbidities.^50,51^

People carrying missense variants were more likely to have anxiety or loss of skills in adulthood, potentially because those with non-missense variants had more severe neurodevelopmental features. Carriers of non-missense variants, in contrast, had a broad range of clinical features. Our reported associations may heighten clinicians’ attention regarding clinical features to remain vigilant for.

Our retrospective perspective from adulthood does not capture the experience of those dying before adulthood. While infections were the principal cause of death, the report of active seizures in the final days of all three emphasizes the importance of seizure control in people with infections and during palliative care.

A history of neonatal seizures appears independent of whether a variant is missense or not, yet both appear to predict lower peak neurodevelopment by CDS and a broader range of comorbidities. We propose recognition of these features as markers of a general prognostic trend but advise against any deterministic interpretation of a formal predictive model based solely on these for any particular person.

Despite five people receiving a molecular diagnosis within 3 years of *CDKL5* being established as a cause of DEE,^2^ most were diagnosed in adolescence. Consequently, if early molecular diagnosis becomes the norm, and if genetically stratified care leads to better management and eventually treatments that mitigate pathogenesis, future cohorts may have a better trajectory. People with more frequent seizures were said to have benefited the most from diagnosis, potentially because of previous reports of treatment outcomes and the emergence of ganaxolone.^36,52–54^

Several methodological issues of our study are notable. In particular, manually coding of highly dimensional clinical data into a formal framework that allows scalable inference has allowed us to maximize the information included in our analyses, despite the challenges of retrospective real-world data, variability in phenotypic screening, and documentation across centers and healthcare systems. We were able to capture a median of 25 independent phenotypes as present and 14 as absent per person, representing 507 harmonized concepts. In contrast, retrospective case series often report only the proportion of individuals known to have had each phenotype.. As previously,^33^ we differentiated between phenotypes that were excluded and those that had not been ascertained in a particular participant. This acknowledges the variability in phenotypic capture from existing clinical data, particularly when collating data from multiple clinical settings and spanning multiple ages, and the inherent uncertainties and inaccuracies that clinicians are used to accommodating. Some phenotypes may not have been reliably documented due to limited relevance to neurological care. Others may be difficult to exclude without investigations that may not have been performed universally, such as osteoporosis. In describing the phenotypic repertoire, we focus on the proportion asserted to have manifest a particular phenotype, as the true value is likely to lie near this lower bound, particularly for neurological phenotypes in the study of neurological disorders. Quantifying the proportion found not have it despite observation and the resulting phenotyping gap allows one to refine the upper limit.^38^ Furthermore, the definite absence of a phenotype can be important because of its functional consequences (for example, the absence of profound intellectual disability or inability to speak), its diagnostic value (the absence of hypsarrhythmia during epileptic spasms is suggestive of CDD),^18^ or its prognostic value (the absence of neonatal seizures (this study) and the absence of cortical visual impairment^18^ are associated with better neurodevelopmental outcomes).

In summary, out study demonstrates the value of critical harmonization and pragmatic retrospective analysis of international real-world data in rare diseases, adding a longitudinal dimension that informs clinical prognosis and management and the design of future clinical trials. The presence of neonatal seizures and classification of the variant may indicate different trajectories, refining prognostication at the time of diagnosis.

## Abbreviations

ASM: anti-seizure medication
CDD: CDKL5 deficiency disorder
CDS: CDKL5 Developmental Score
DEE: developmental and epileptic encephalopathy
EEG: electroencephalogram
FET: Fisher’s exact test
HPO: Human Phenotype Ontology
ICDD: International CDKL5 Disorder Database
*minP*: minP stepdown *p*-value
MRI: magnetic resonance imaging
OR: odds ratio
RR: relative risk
WRST: Wilcoxon rank sum test with continuity correction
95%CI: 95% confidence interval

## Data Availability

All data produced in the present study are available upon reasonable request to the authors

## Acknowledgements

The authors wish to thank the people with CDD and their caregivers who consented to use of their clinical data for this study. Data were collected with the aid of the Loulou Foundation, the CDKL5 Alliance, including various national CDKL5 advocacy groups, the International CDKL5 Disorder Database, and the EpiCARE platform for collaborative genetic research.

