## Supplemental Fig 1-16 for "The natural history of CDKL5 deficiency disorder into adulthood"

### Slide 1
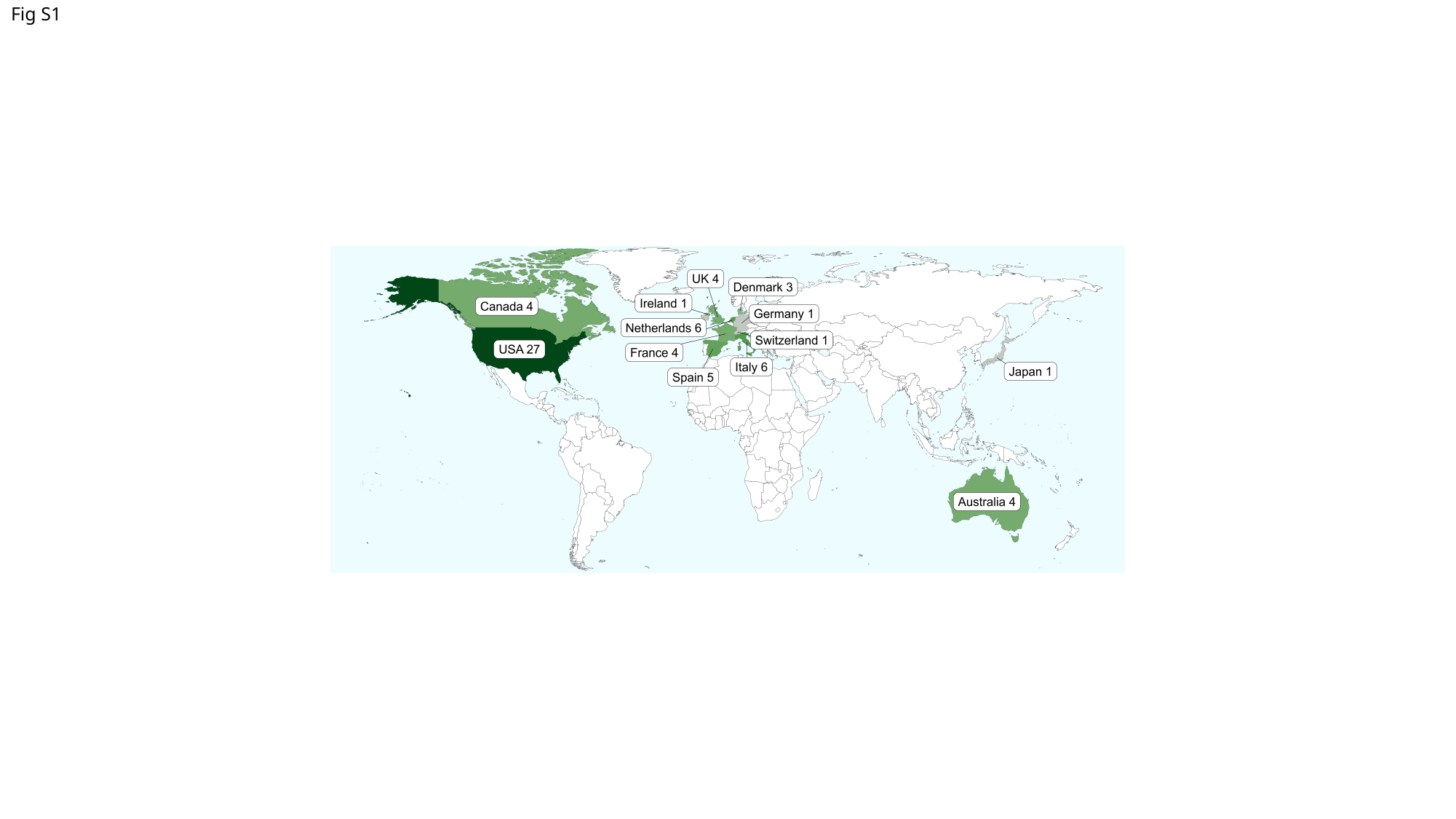

#### Fig S1

### Slide 2
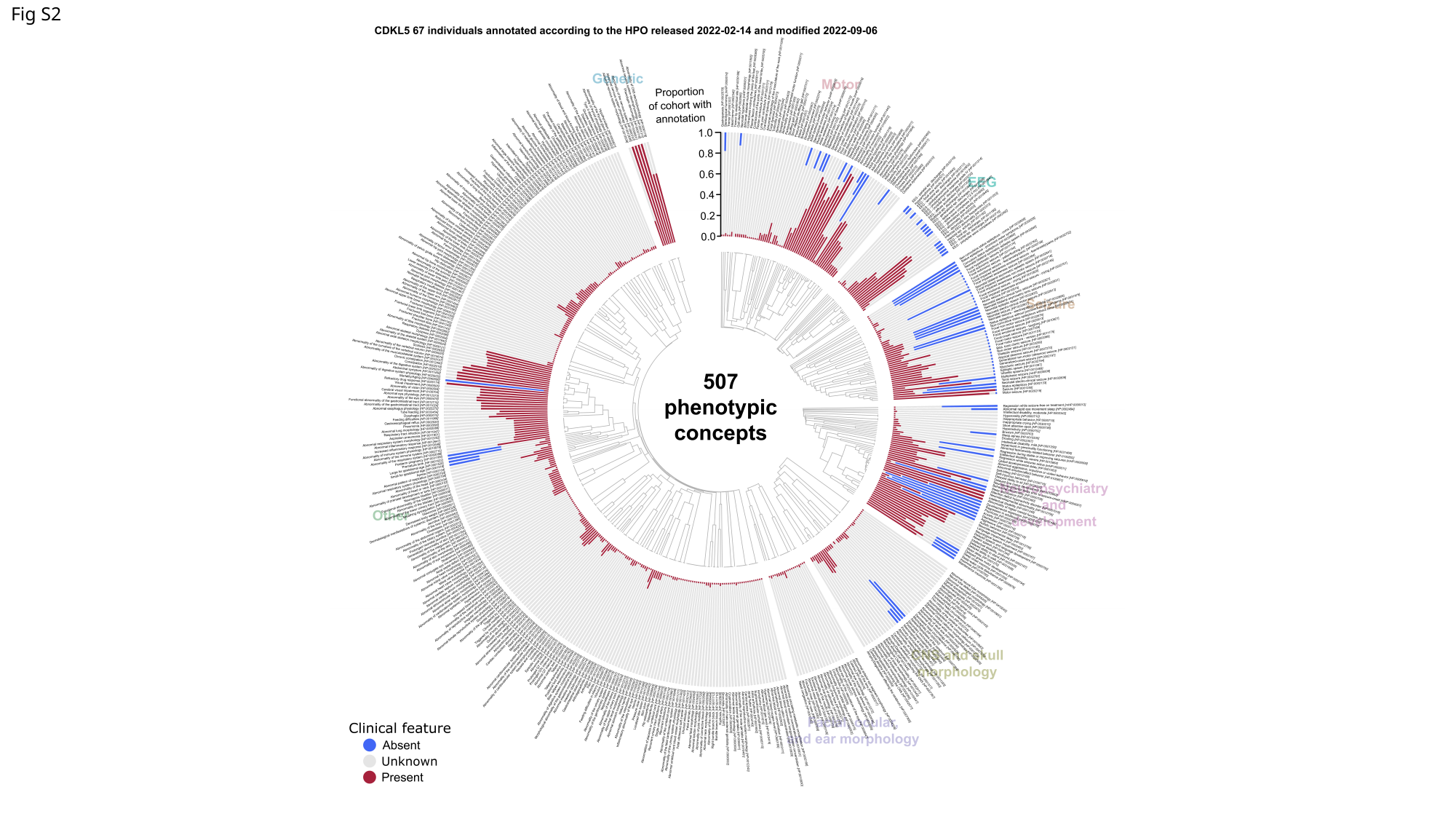

Fig S2

### Slide 3
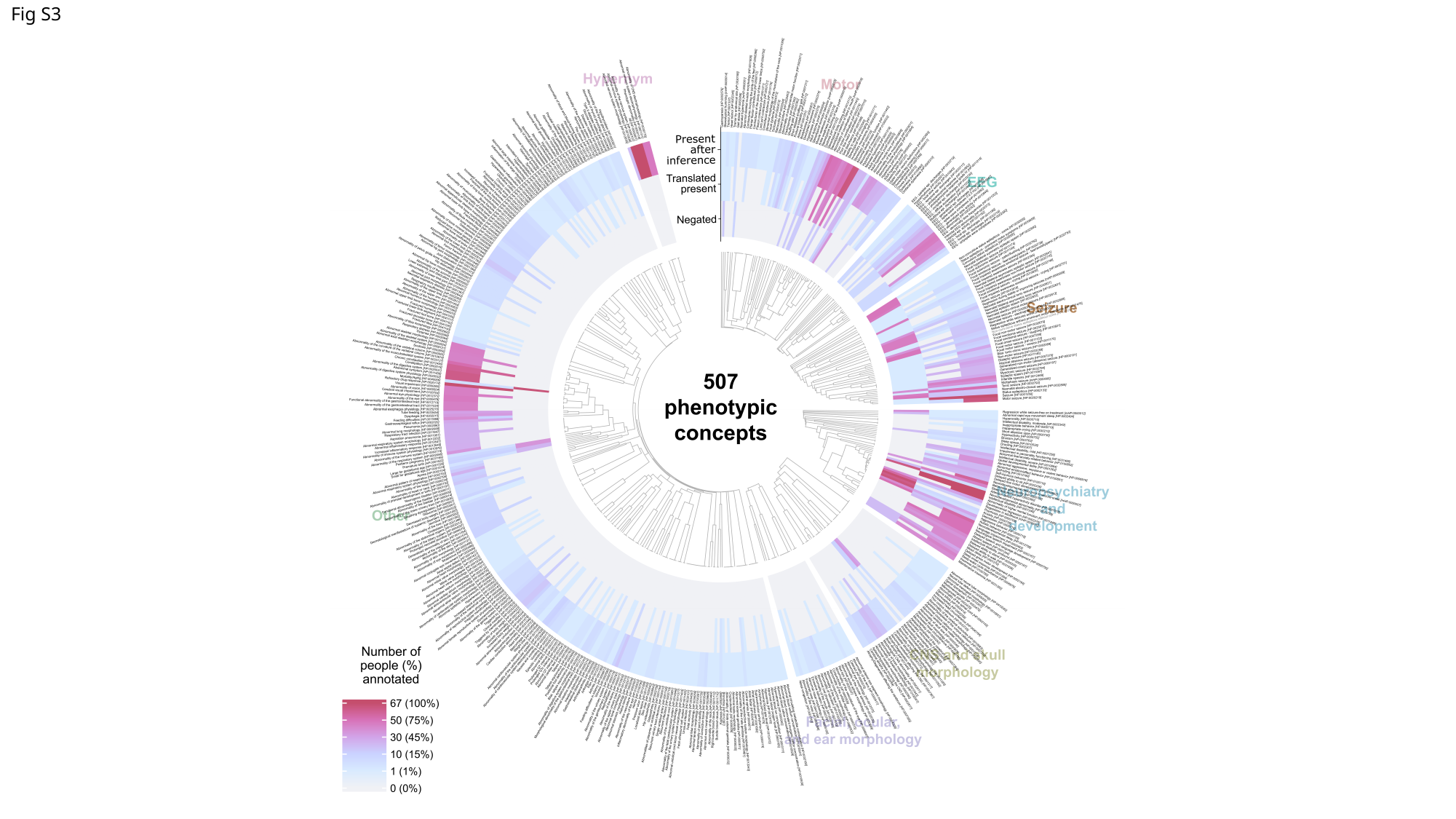

Fig S3

### Slide 4
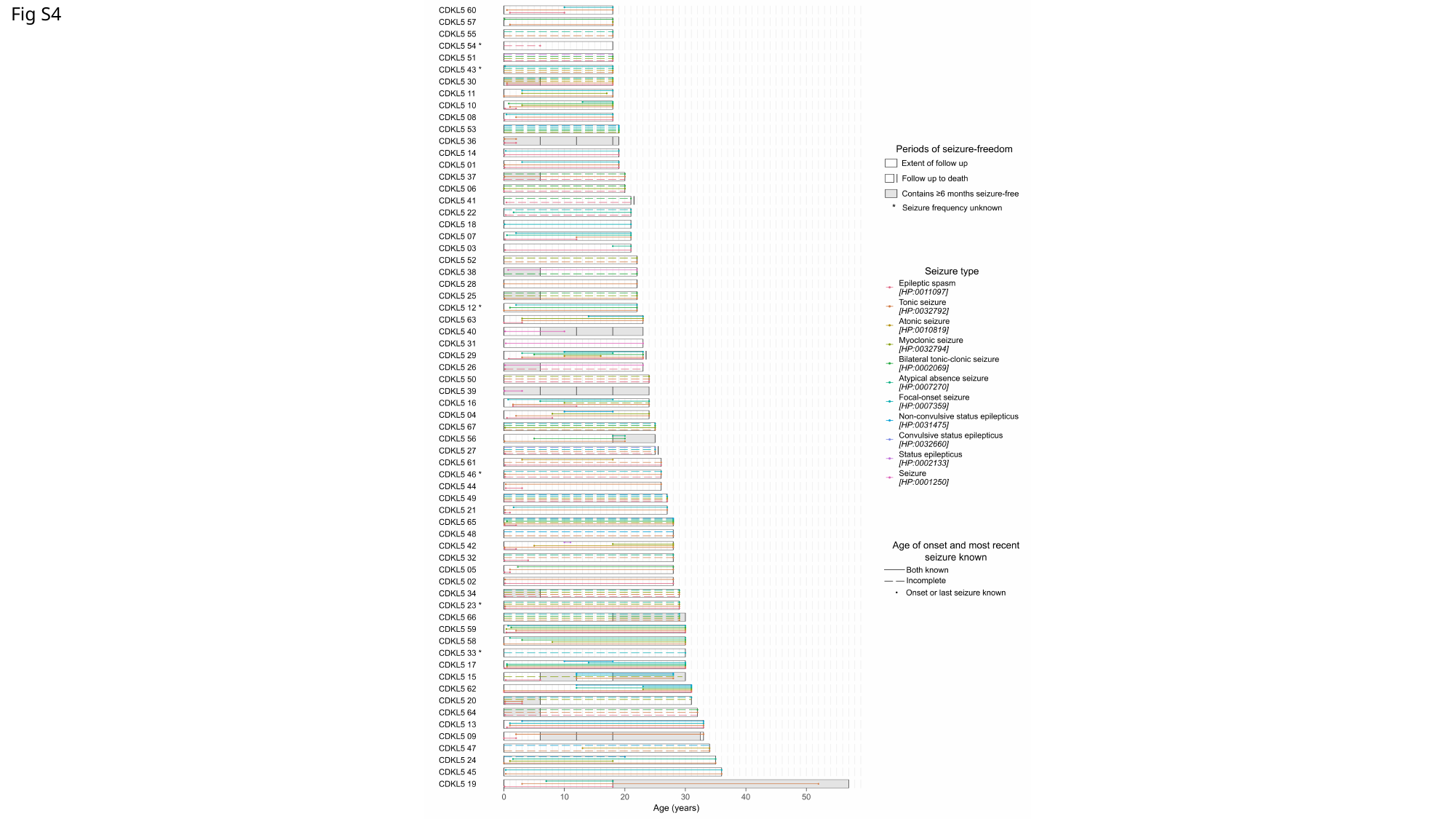

Fig S4

### Slide 5
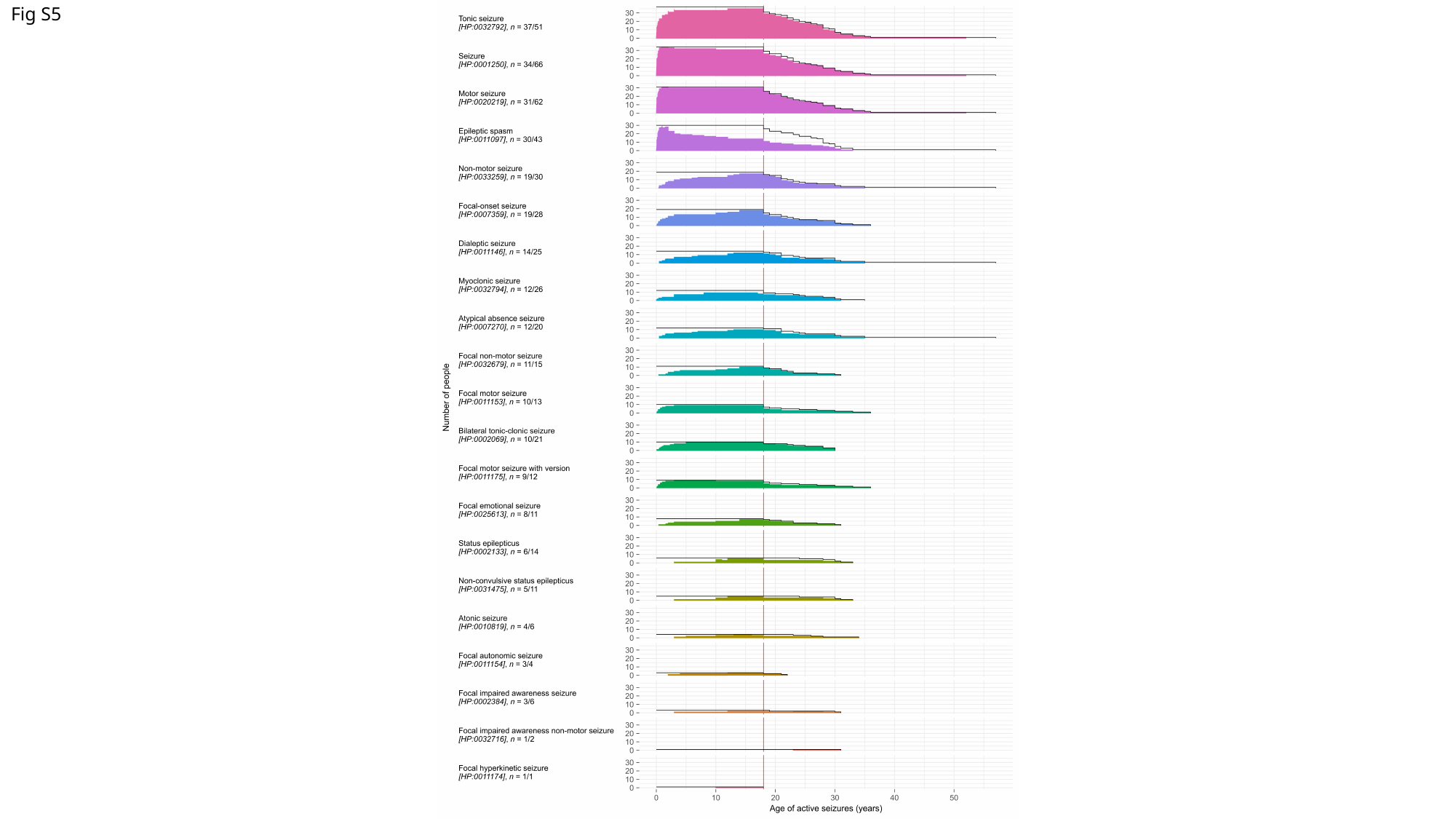

Fig S5

### Slide 6
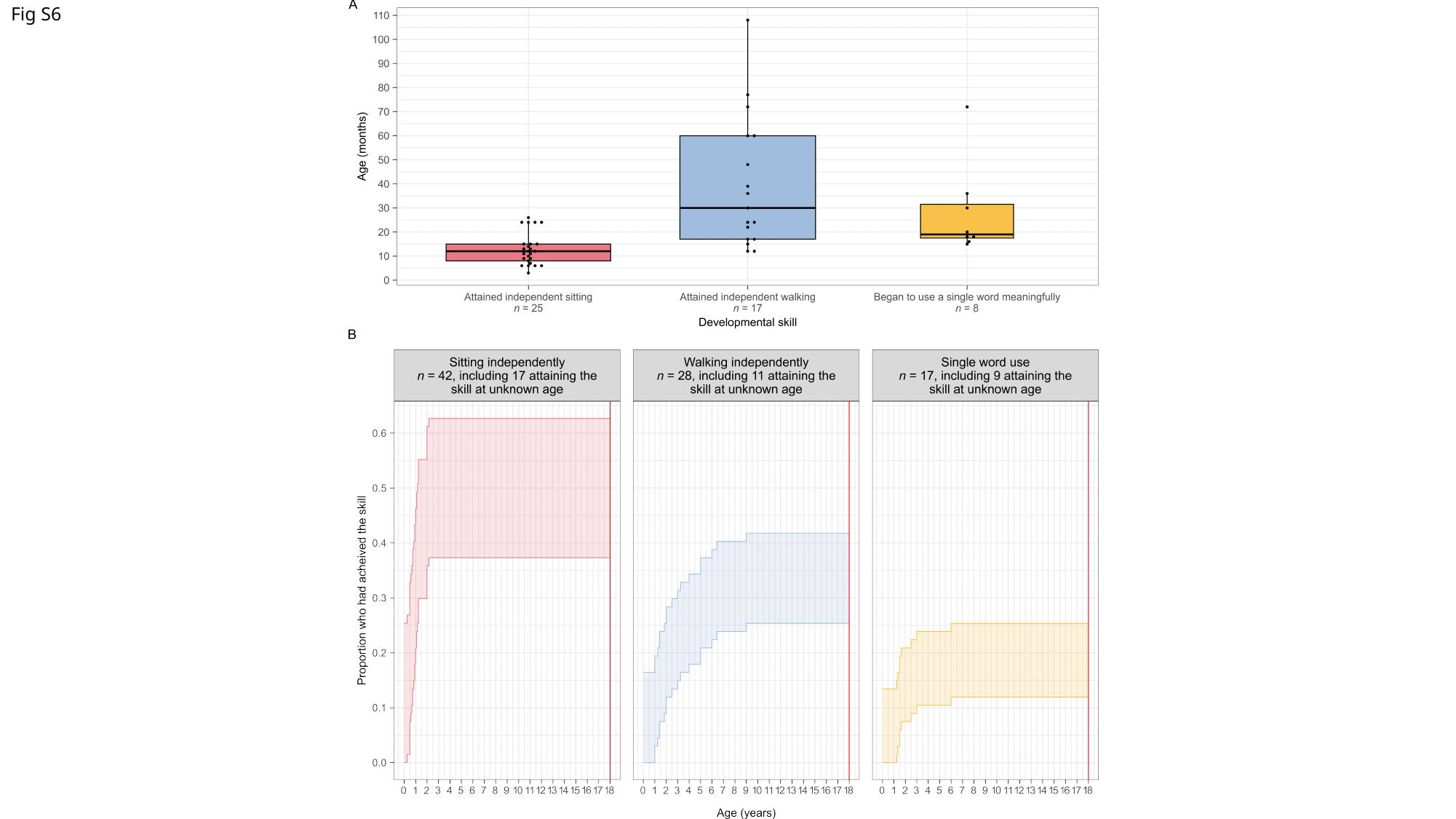

Fig S6

### Slide 7
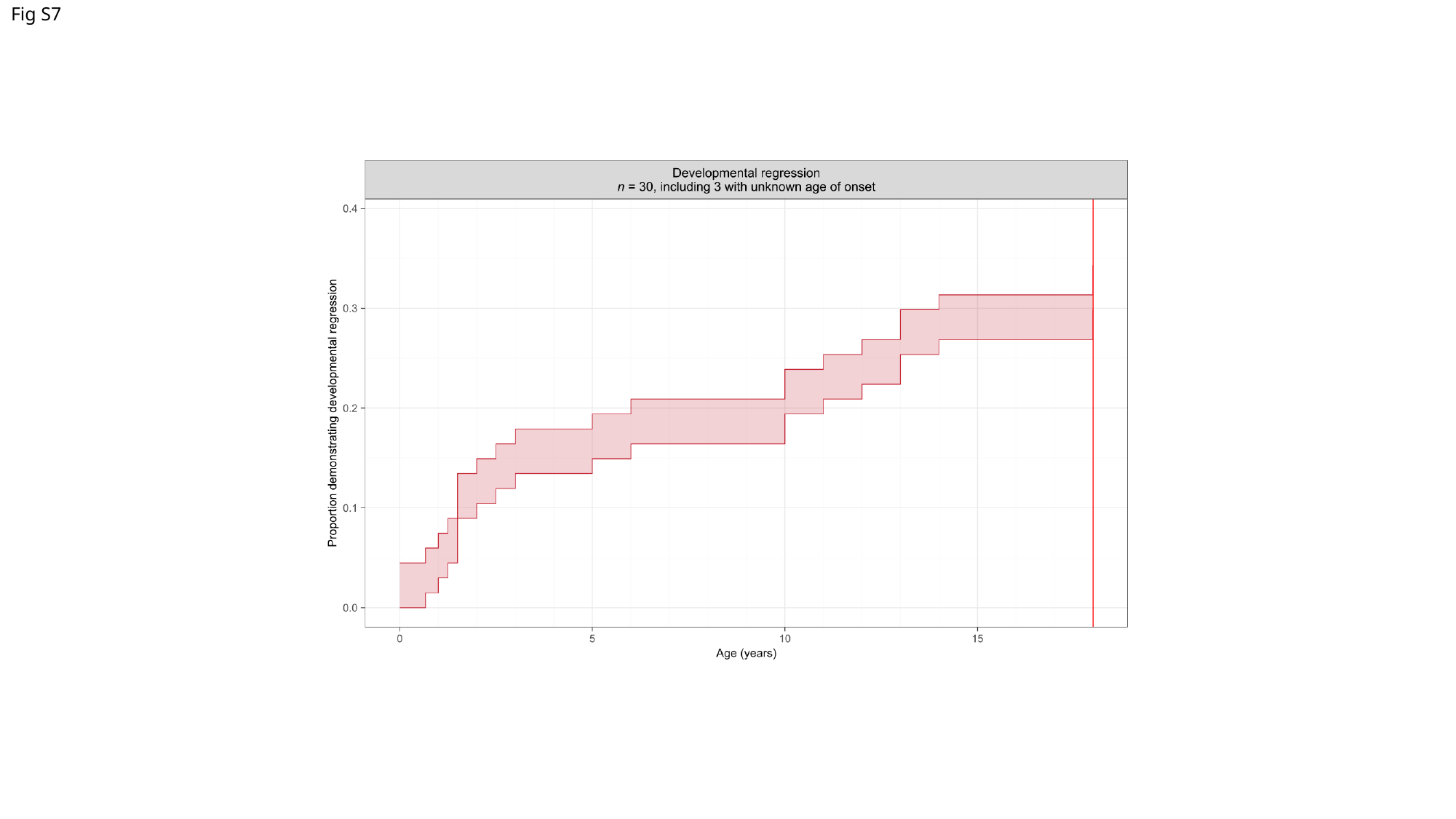

Fig S7

### Slide 8
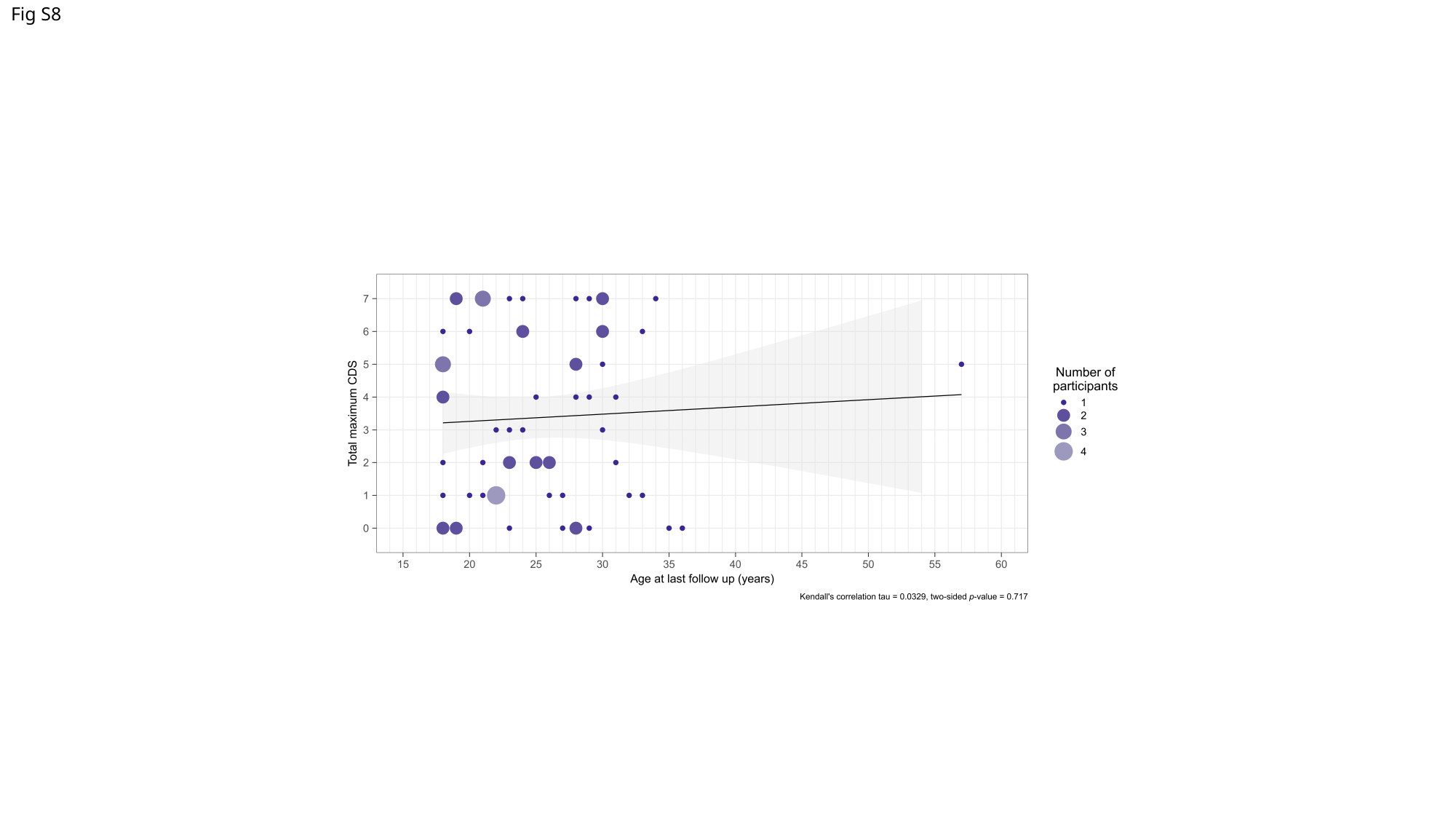

Fig S8

### Slide 9
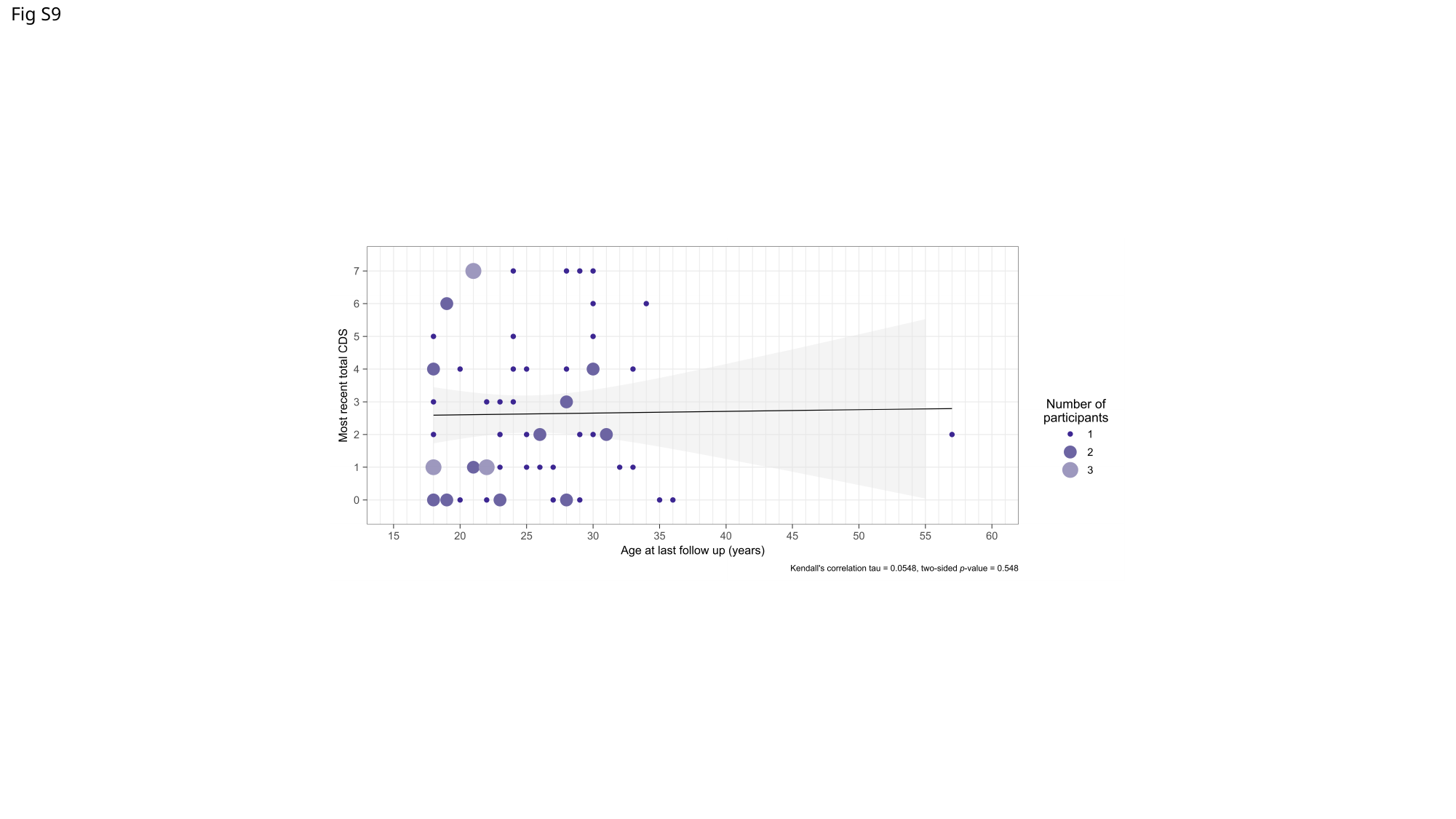

Fig S9

### Slide 10
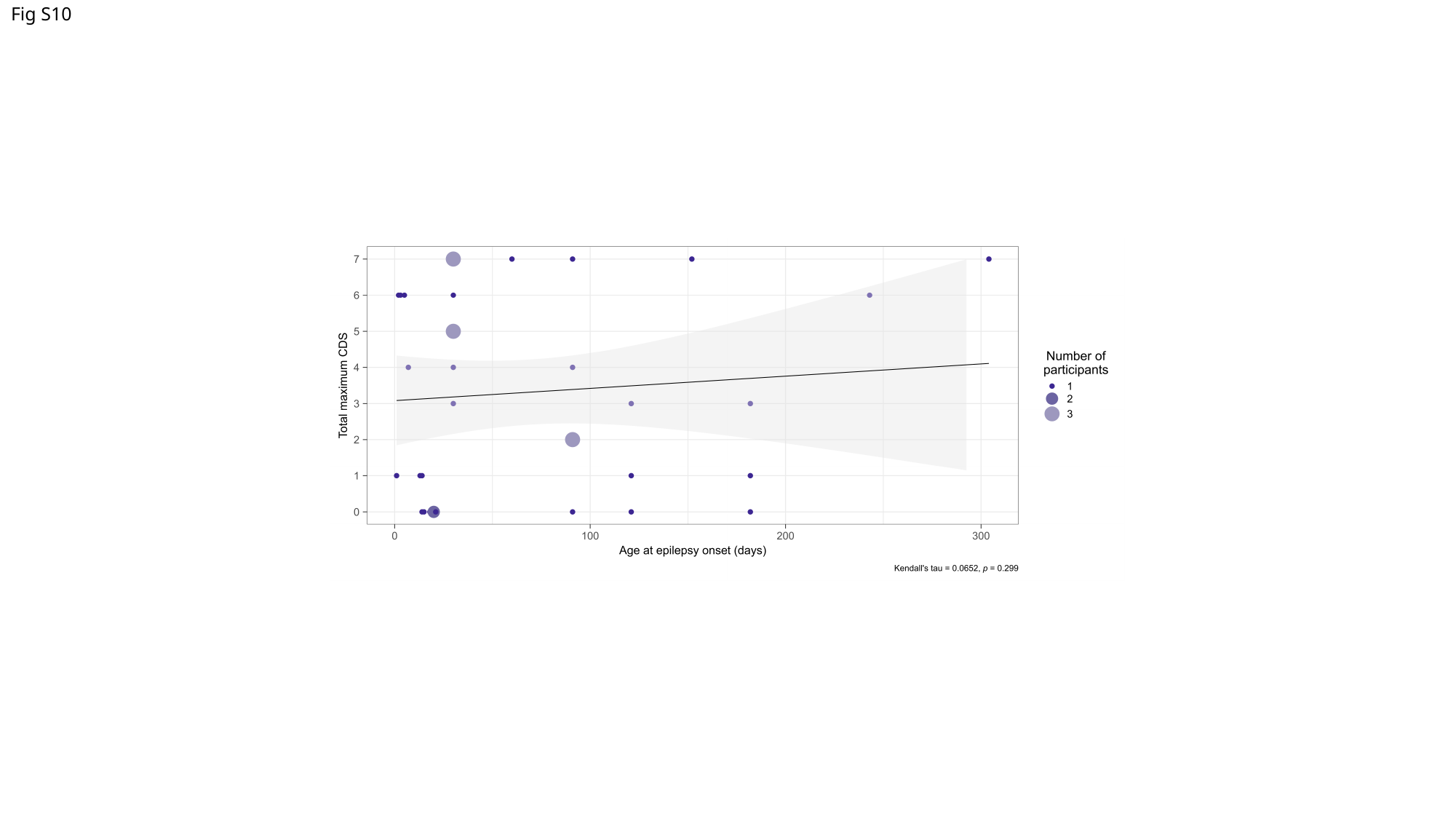

Fig S10

### Slide 11
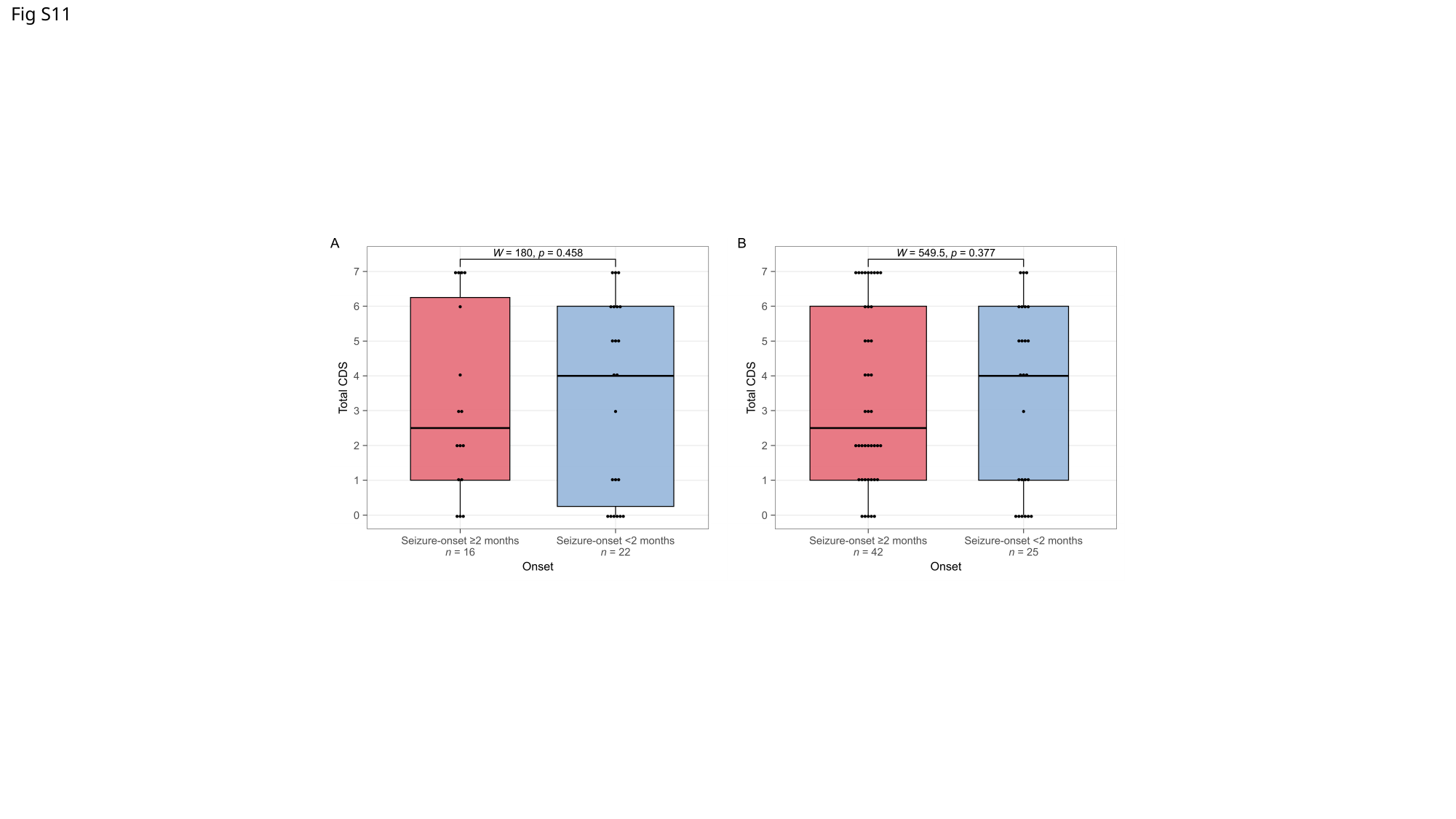

Fig S11

### Slide 12
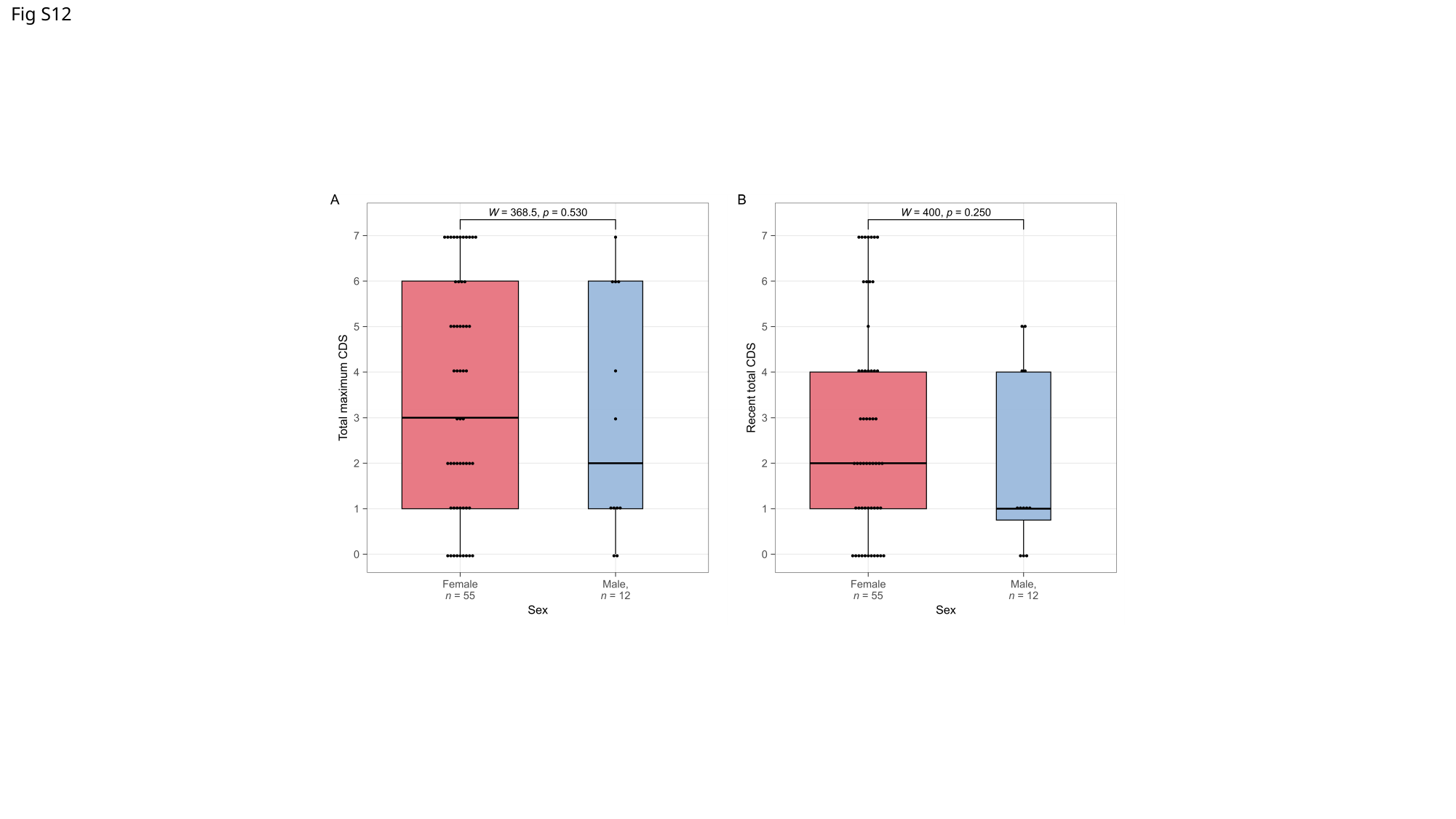

Fig S12

### Slide 13
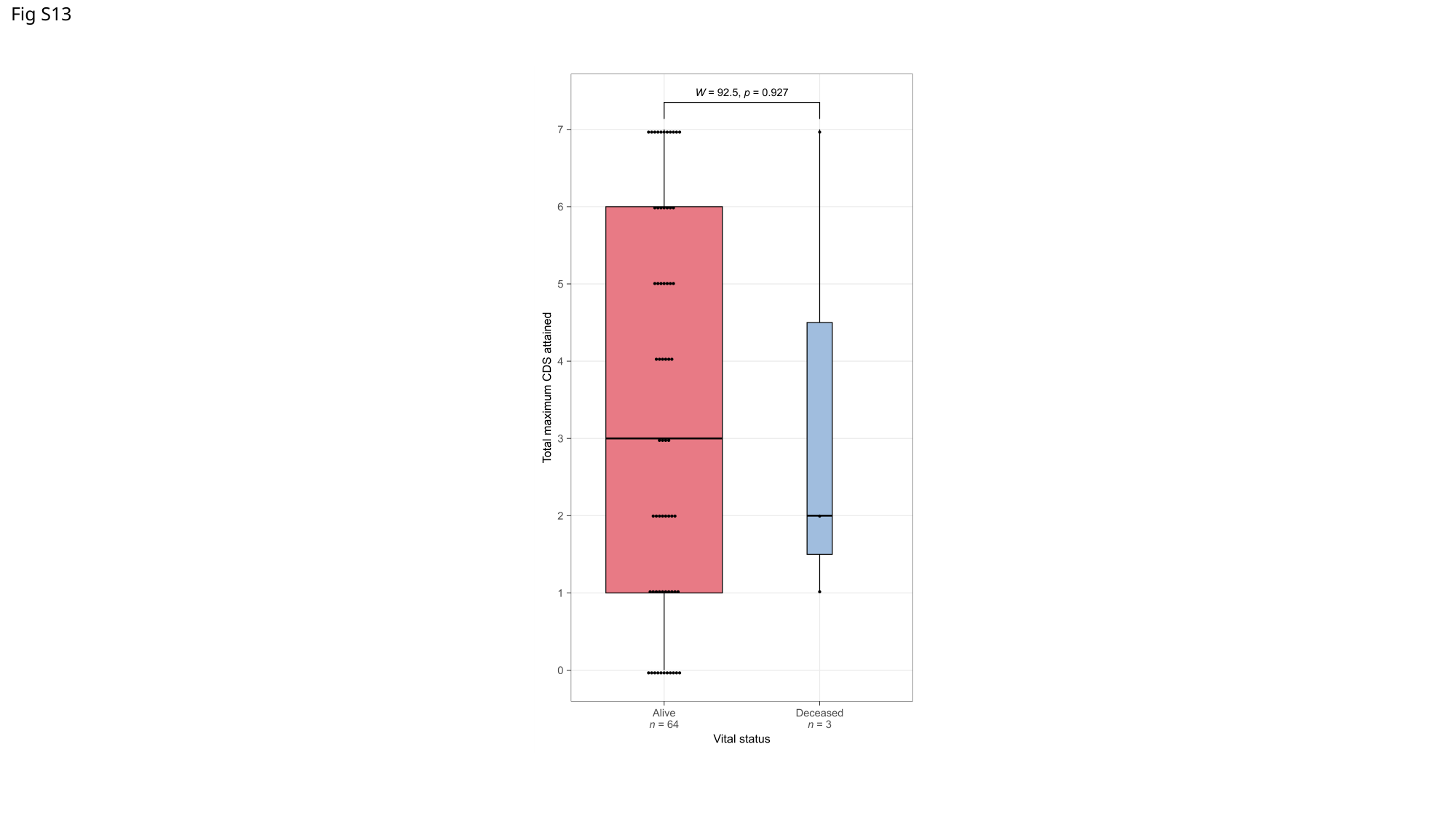

Fig S13

### Slide 14
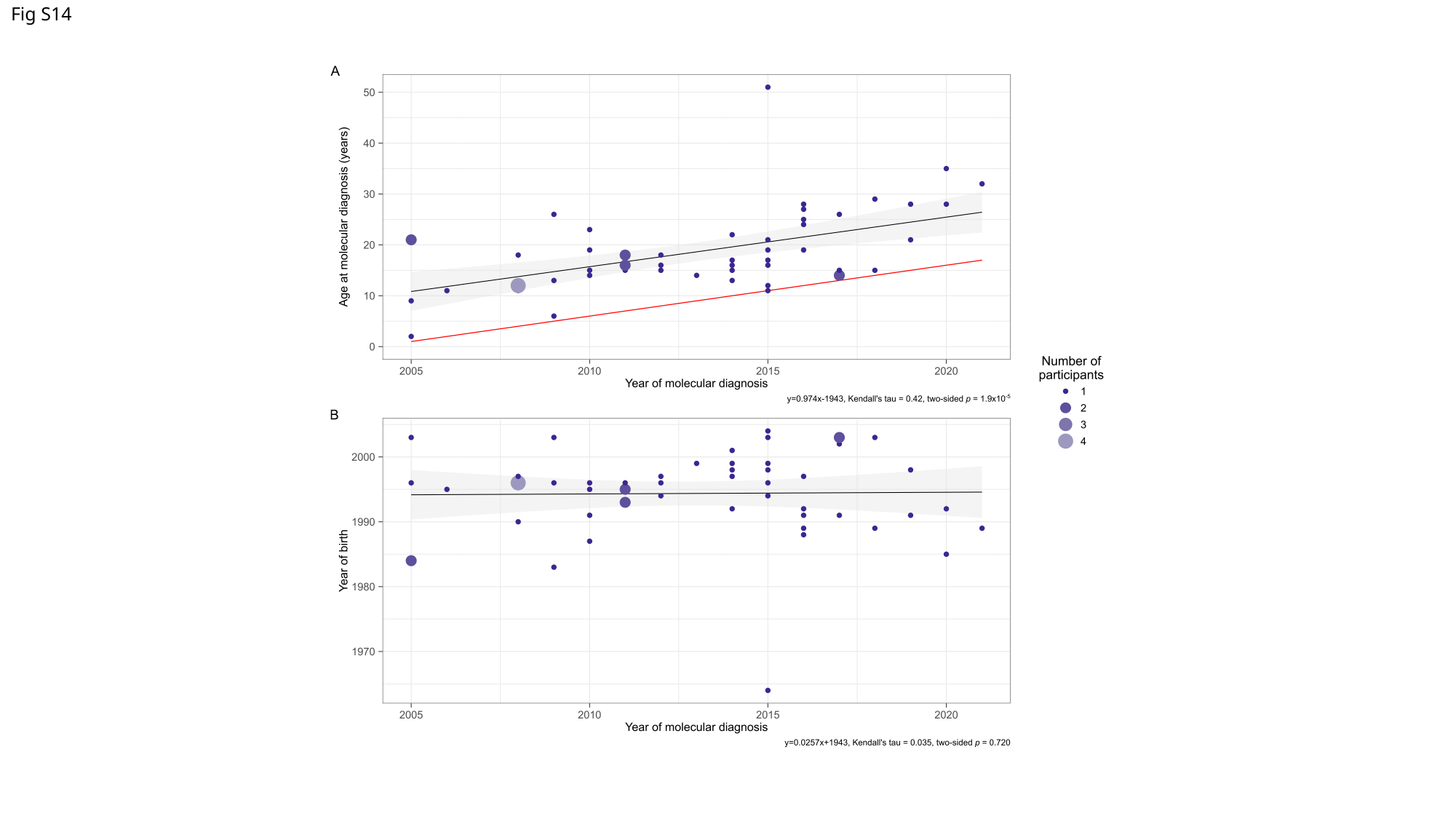

Fig S14

### Slide 15
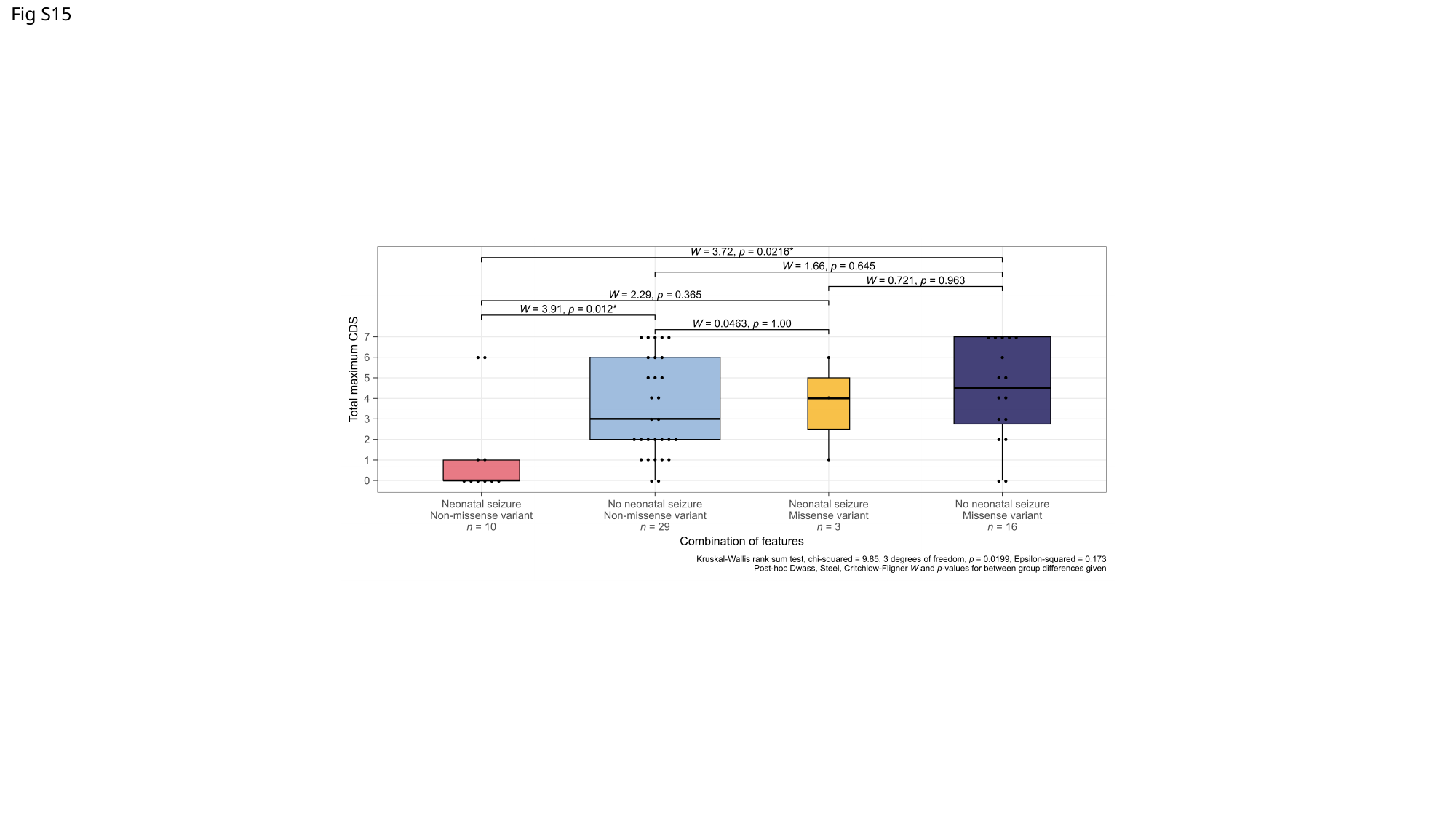

Fig S15

### Slide 16
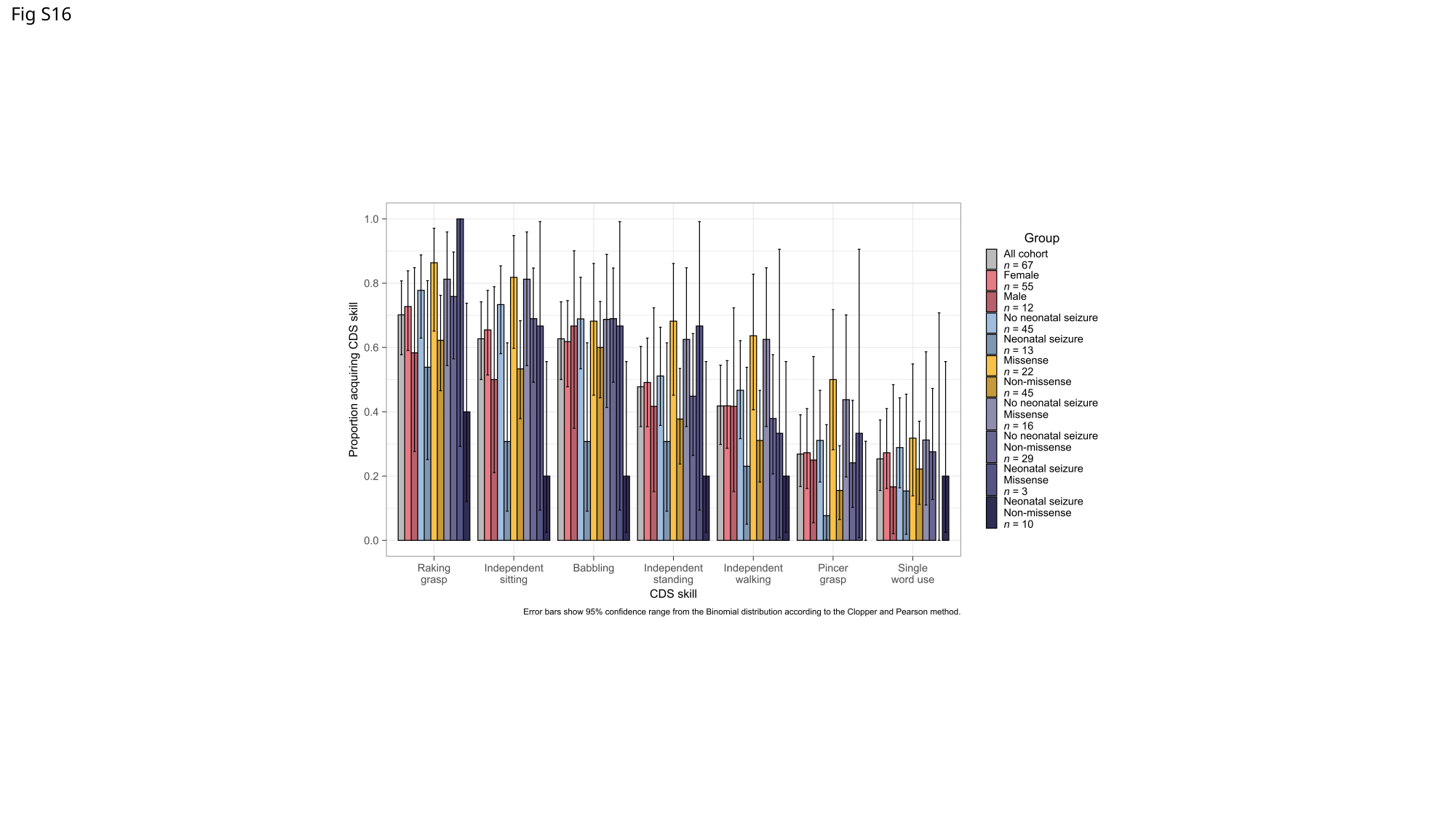

Fig S16
